# Analytical and Clinical Validation of Direct Detection of Antimicrobial Resistance Markers by Plasma Microbial Cell-free DNA Sequencing

**DOI:** 10.1101/2024.03.14.24303933

**Authors:** Fred C. Christians, Jamilla Akhund-Zade, Kristin Jarman, Shivkumar Venkatasubrahmanyam, Nicholas Noll, Timothy A. Blauwkamp, Sivan Bercovici, Aga Zielinska, Amy L. Carr, Arryn Craney, Matthew Pike, John Joseph Farrell, Sanjeet Dadwal, James B. Wood, Efrat Matkovich, Staci McAdams, Frederick S. Nolte

**Affiliations:** Karius, Redwood City, CA; AdventHealth Orlando, Orlando, FL; Orlando Health, Orlando, FL; Carle Foundation Hospital, Urbana, IL; OSF Healthcare, Peoria, IL; City of Hope National Medical Center, Duarte. CA; Indiana University School of Medicine,Indianapolis, IN

## Abstract

Sequencing of plasma microbial cell-free DNA (mcfDNA) has gained increased acceptance as a valuable adjunct to standard-of-care testing for diagnosis of infections throughout the body. Here we report the analytical and clinical validation of a novel application of mcfDNA sequencing, the non-invasive detection of seven common antimicrobial resistance (AMR) genetic markers in 18 important pathogens with potential to harbor these markers. The AMR markers include SCC*mec*, *mecA* and *mecC* for methicillin, *vanA* and *vanB* for vancomycin, *bla*_CTX-M_ for oxyimino-cephalosporin and aztreonam, and *bla*_KPC_ for carbapenem resistance. The AMR markers are computationally linked to the pathogens detected, using a statistical model based on observed AMR gene and pathogen abundances. Analytical validation showed high reproducibility (100%), inclusivity (54 to100%), and exclusivity (100%), with limits of detection ranging from 425 to 6,107 pathogen mcfDNA molecules/μL for the different markers. Clinical accuracy was assessed with 115 unique plasma samples from patients at 7 study sites with concordant culture results for 12/18 (66.7%) target bacteria from a variety of specimen types and correlated with available phenotypic antimicrobial susceptibility test results and genotypic results when available. The positive percent agreement (PPA), negative percent agreement (NPA), overall percent agreement (OPA), and diagnostic yield (DY) were estimated for each AMR marker. The results for the combination of SCC*mec* and *mecA* for staphylococci were PPA 19/20 (95.0%), NPA 21/22 (95.4%), OPA 40/42 (95.2%), DY 42/60 (70.0%); *vanA* for enterococci were PPA 3/3 (100%), NPA 2/2 (100%), OPA5/5 (100%), DY 5/6 (83.3%); *bla*_CTX-M_ for gram-negative bacilli were PPA 5/6 (83.3%), NPA 29/29 (100%), OPA34/35 (97.1%), DY 35/49 (71.4%); and *bla*_KPC_ for gram-negative bacilli were PPA 0/2 (0%), NPA: 23/23 (100%), OPA23/25 (92.3%), DY 25/44 (56.8%). The addition of AMR capability to plasma mcfDNA sequencing should provide clinicians with an effective new culture-independent tool for optimization of therapy.

## INTRODUCTION

Antimicrobial resistance (AMR) is one of the single greatest concerns for human health globally (1). The detection of AMR is traditionally achieved by phenotypic testing. However, a limitation of the phenotypic approach is its time requirement and low yield of cultures that may prevent or delay optimal antimicrobial therapy and result in poor health outcomes. As most clinically important AMR has a genetic basis, molecular methods to directly detect resistance without the need for culture are attractive. However, the molecular detection of resistance determinants also poses significant challenges, foremost of which is the genetic complexity of AMR. Resistance can arise due to the acquisition of resistance determinants (e.g., β-lactamase genes) but can also reflect changes in gene expression (e.g., efflux pumps and porins) or the acquisition of mutations (e.g., fluoroquinolone target mutations). Additionally, the detection of a resistance gene may not necessarily imply phenotypic resistance if the gene is expressed at low levels (2).

Multiplex-PCR syndromic panels that identify common pathogens and AMR determinants within a few hours from positive blood cultures or directly from some specimen types such as bronchoalveolar lavage (BAL), sputum, and joint fluid are widely deployed in clinical microbiology laboratories (3). However, these panels can only identify a limited number of pathogens and resistance determinants. In contrast, whole-genome sequencing (WGS) is the most comprehensive molecular method to detect AMR because it allows the simultaneous detection of resistance to numerous antimicrobial classes (4, 5). WGS is particularly attractive for its potential to identify novel or variant resistance determinants that might not be detected by targeted assays and for its utility as an epidemiologic tool to assess transmission dynamics of pathogenic multidrug-resistant organisms during an outbreak. WGS can be used either to detect AMR in culture isolates or directly from clinical specimens. However, WGS is predicated on a detailed understanding of the relationship between genotype and resistance phenotype that is presently incomplete and may require advanced machine learning algorithms to improve the ability to accurately predict AMR from WGS data (6, 7). Resistance mechanisms relating to the presence or absence of specific genes or well-characterized target site mutations will be more straightforward to interpret than those relating to alterations in gene expression.

In a systematic review of the reported diagnostic accuracy of 13 metagenomic tests, AMR prediction and comparison to phenotypic results were undertaken in four (4). Overall percent agreement was 88%, and very major and major error rates were 24% and 5%, respectively. The Food and Drug Administration (FDA) uses a very major error rate of <1.5% and a major error rate of <3% as acceptable criteria when comparing different test antimicrobial susceptibility testing methods (8). For most bacterial species the major limitations to widespread adoption for WGS-based antimicrobial susceptibility testing (AST) in clinical laboratories remain the high cost and limited speed of inferring antimicrobial susceptibility from WGS data as well as the dependency on previous culture because analysis directly on specimens remains challenging. For most bacterial species there is insufficient evidence to support the use of WGS-inferred AST to guide clinical decision making (9).

Metagenomic sequencing of plasma microbial cell-free DNA (mcfDNA) has gained increased acceptance as a valuable adjunct to standard-of-care testing for the diagnosis of infections throughout the body. The Karius Test® is an analytically and clinically validated mcfDNA sequencing test, commercially available since December 2016 as a laboratory developed test. The test identifies and quantitates mcfDNA in plasma for >1,000 pathogens including bacteria, DNA viruses, fungi, and parasites within a clinically relevant time frame, with a median time from specimen collection to report of 2.6 days (10). The analytical and clinical validation of the test has been previously reported (11). Since being clinically available, substantial evidence has emerged documenting its clinical impact by potentially reducing invasive procedures (12), reducing time to specific etiologic diagnosis and increasing diagnostic yield as compared with standard of care microbiological testing (11, 12, 13, 14, 15), and optimizing antimicrobial therapy (16, 17, 18, 19, 20) in a variety of patient populations.

The first AMR capability incorporated into the Karius Test became available in April 2022 was a computational enhancement for samples with *Staphylococcus aureus* calls to determine the presence or absence of the SCC*mec* element, which harbors the *mecA* or *mecC* resistance gene (21). While the analytical and clinical accuracy of this capability is very high and the results are available on the same day as identification of the potential pathogen, the diagnostic yield is relatively low, at approximately 50%, because there is no enrichment for SCC*mec*.

To increase the diagnostic yield of AMR testing for staphylococci and to add additional AMR markers to the test, we developed a more sensitive assay that directly targets a panel of 6 additional common AMR genes in 18 selected bacteria encoding resistance to methicillin (*mec*A and *mec*C), vancomycin (*van*A and *van*B), oxyimino-cephalosporins and aztreonam (*bla*_CTX-M_), and carbapenems (*bla*_KPC_). Consistent with good laboratory practice, all the AMR markers are linked to the organism detected, and the report provides guidance to assist in optimization of antimicrobial management (22).

This new capability represents a novel application of plasma mcfDNA sequencing to detect AMR genes by enriching for highly degraded, ultra-rare target fragments in plasma prior to sequencing without the need for a positive culture or larger genomic fragments. Here we report the analytical and clinical validation of mcfDNA sequencing for detection of AMR genetic markers that were added to the Karius Test workflow. The test with expanded capabilities for prediction of AMR was commercially launched in September 2023.

## MATERIAL AND METHODS

### Karius Test with AMR

mcfDNA was extracted from 250 μL of plasma, converted to DNA libraries, and sequenced on an Illumina NextSeq®500 or NovaSeq® at a CAP-accredited and CLIA-certified laboratory according to previously validated methods (Karius, Redwood City, CA). Any of the over 1,000 organisms included in the Karius clinical reportable range found to be present above a predefined statistical threshold were reported as previously described. The quantity for each organism identified was expressed in molecules per microliter (MPM), representing the number of mcfDNA molecules from the reported microorganism present per microliter of plasma (absolute quantity). For monitoring the yield of the Karius Test laboratory workflow, whole assay internal control (WINC) molecules are spiked into each sample at a known concentration as a control for sequencing depth or human background levels. A quality control minimum of 25,000 unique WINC molecules was set for reporting microorganism absolute quantities in MPM (11).

For samples in which *S. aureus* is reported, the Karius Test analytical pipeline further reports methicillin-resistant *S. aureus* (MRSA) when there is strong evidence consistent with the presence of a *mecA* or *mecC* containing SCC*mec* cassette, methicillin-susceptible *S. aureus* (MSSA) when there is strong evidence for the lack of an SCC*mec* cassette, or indeterminate when the evidence is ambiguous or insufficient. Broadly, the determination is based on the number of sequencing reads that align to a collection of SCC*mec* alleles (44 SCC*mec* variants and 4 non-*mec* SCC variants), a 20-70 kb genetic element known to harbor the *mecA* or *mecC* genes and integrated into the bacterial chromosome (23). The number of sequencing reads is compared to the number of reads mapping to the *S. aureus* genome and a statistical model is used to determine the likelihood of either the presence or absence of methicillin resistance. To reduce false-positive MRSA calls, indeterminate is also reported if the SCC in the clinical sample is likely to be a non-*mec* SCC or if SCC fragments in the clinical sample may have originated from a microbe other than *S. aureus*, as determined by comparing the abundances of a set of interfering species, known to harbor sequences highly homologous to SCC*mec,* to that of *S. aureus*.

The AMR gene detection workflow (Fig. 1) is triggered by Karius Test results that reported one or more of the 18 targeted bacteria (Table 1) provided that additional requirements were met: sufficient plasma volume remaining (>500 μL); for *bla*_CTX-M_ and *bla*_KPC_ testing, the target bacterial species is reported at >1000 MPM; for *S. aureus*, the SCC*mec* caller reports “indeterminate”; and for *S. aureus* with MPM < 300, the abundance of interfering coagulase-negative staphylococci is <5 MPM.

**Fig. 1.**
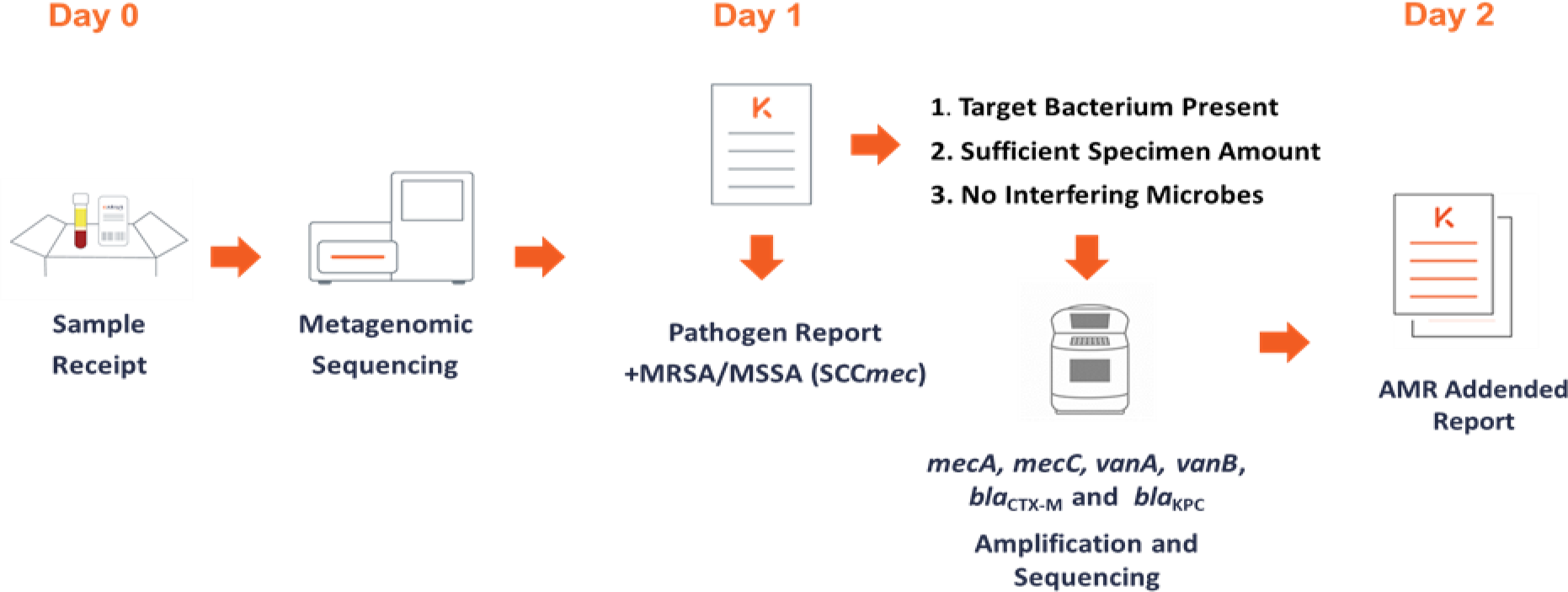
Overview of Karius Test AMR workflow. See text for more details.

**Table 1.**
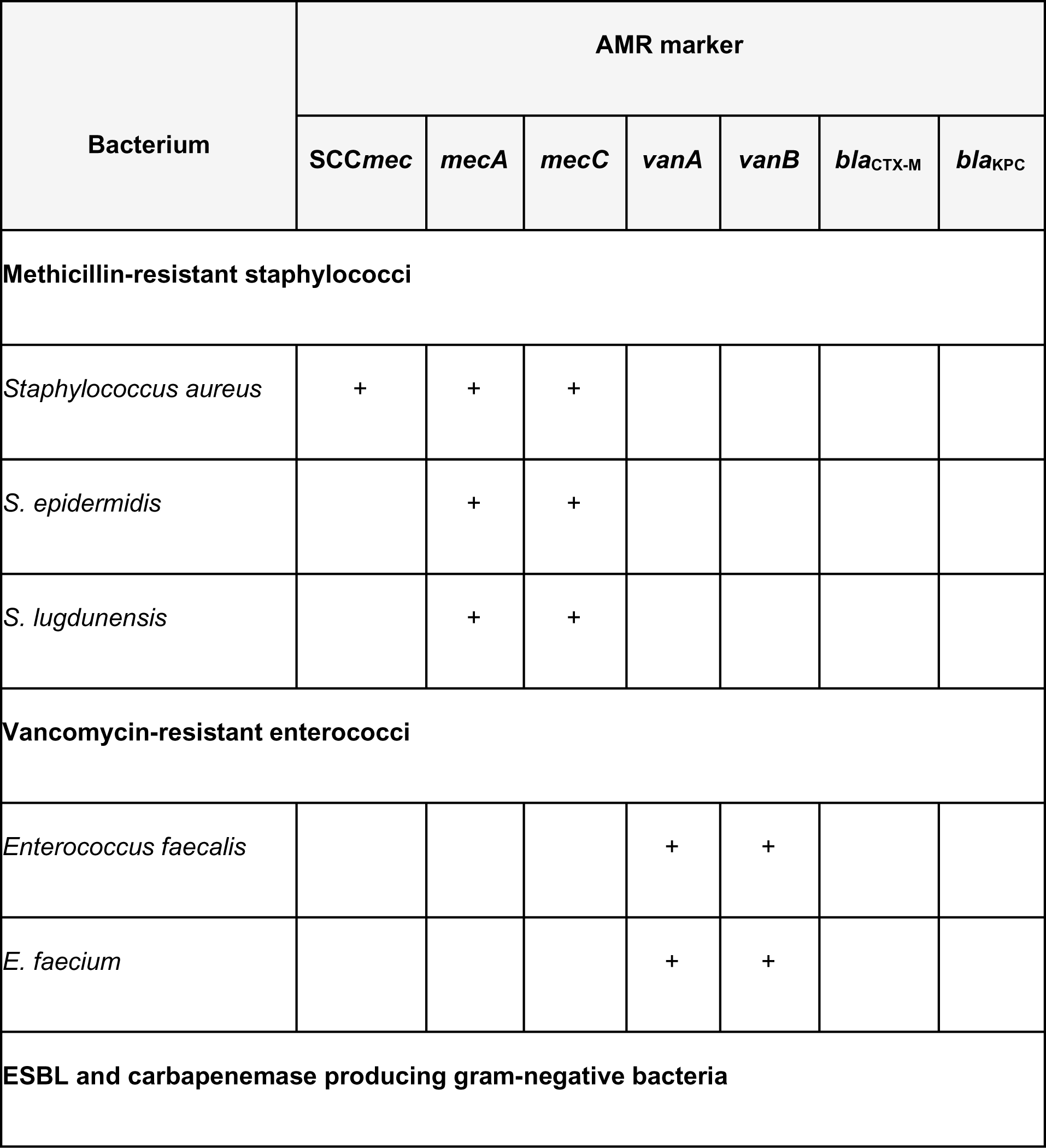
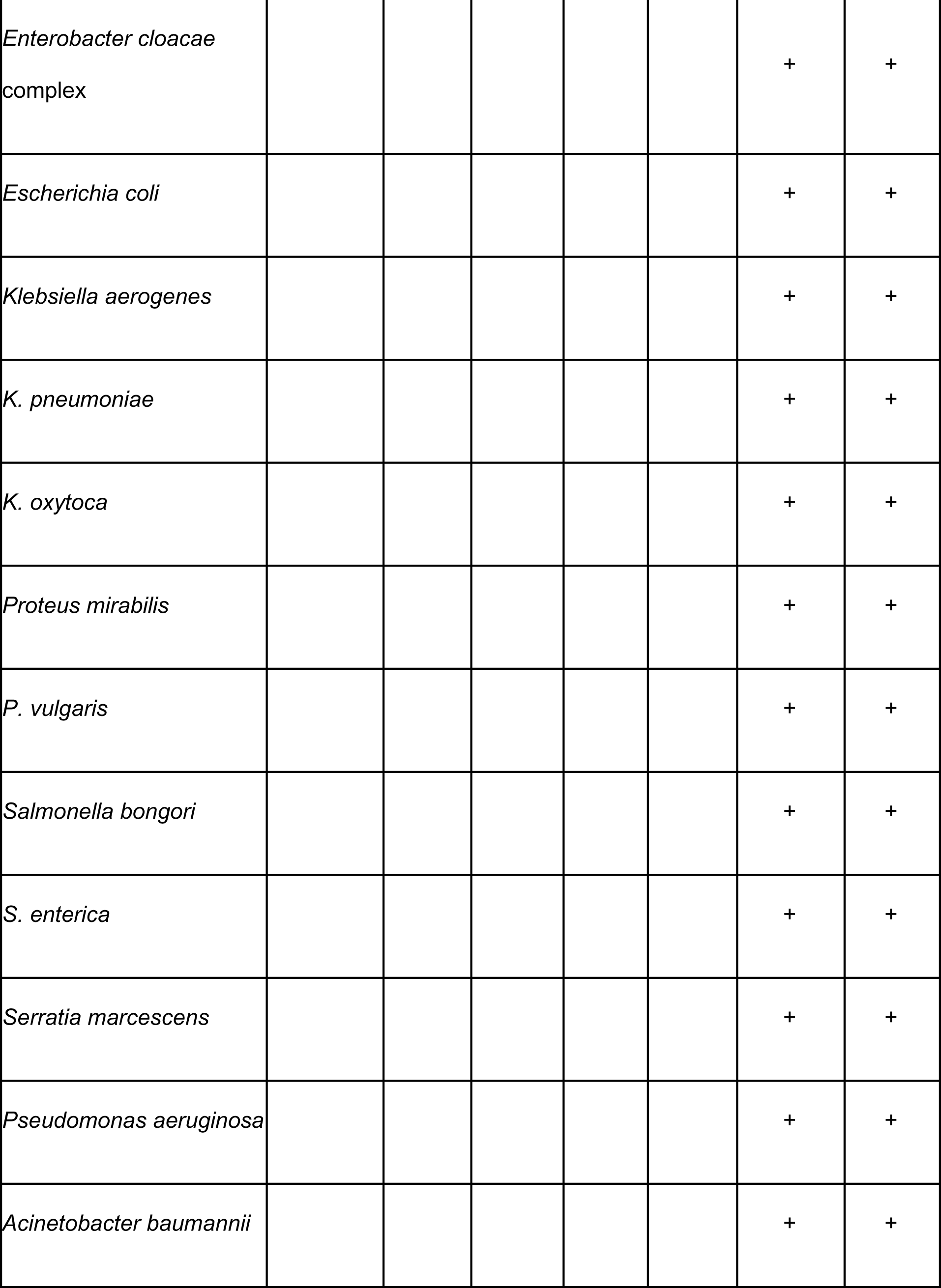
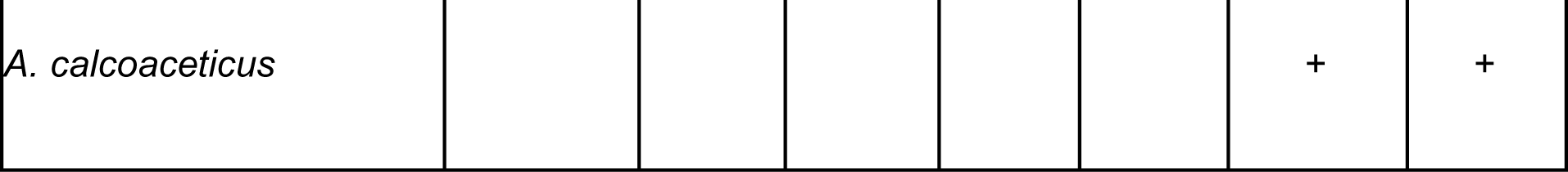
Target Bacteria and associated AMR markers.

| Bacterium | AMR marker |  |  |  |  |  |  |
| --- | --- | --- | --- | --- | --- | --- | --- |
|  | SCC<sub>mec</sub> | mecA | mecC | vanA | vanB | bla<sub>CTX-M</sub> | bla<sub>KPC</sub> |
| **Methicillin-resistant staphylococci** |  |  |  |  |  |  |  |
| *Staphylococcus aureus* | + | + | + |  |  |  |  |
| *S. epidermidis* |  | + | + |  |  |  |  |
| *S. lugdunensis* |  | + | + |  |  |  |  |
| **Vancomycin-resistant enterococci** |  |  |  |  |  |  |  |
| *Enterococcus faecalis* |  |  |  | + | + |  |  |
| *E. faecium* |  |  |  | + | + |  |  |
| **ESBL and carbapenemase producing gram-negative bacteria** |  |  |  |  |  |  |  |
| *Enterobacter cloacae*<br>complex |  |  |  |  |  | + | + |
| *Escherichia coli* |  |  |  |  |  | + | + |
| *Klebsiella aerogenes* |  |  |  |  |  | + | + |
| *K. pneumoniae* |  |  |  |  |  | + | + |
| *K. oxytoca* |  |  |  |  |  | + | + |
| *Proteus mirabilis* |  |  |  |  |  | + | + |
| *P. vulgaris* |  |  |  |  |  | + | + |
| *Salmonella bongori* |  |  |  |  |  | + | + |
| *S. enterica* |  |  |  |  |  | + | + |
| *Serratia marcescens* |  |  |  |  |  | + | + |
| *Pseudomonas aeruginosa* |  |  |  |  |  | + | + |
| *Acinetobacter baumannii* |  |  |  |  |  | + | + |
| *A. calcoaceticus* |  |  |  |  |  | + | + |

In the AMR wet-lab workflow, 500 µL of plasma was spiked with two synthetic control DNA molecules, an ID spike and an AMR WINC. mcfDNA was extracted, followed by targeted amplification using more than 300 primers to amplify mcfDNA fragments from *mecA*, *mecC*, *vanA*, *vanB*, *bla*_CTX-M_, *bla*_KPC_, several species-specific housekeeping genes, and the synthetic control molecules. Following sequencing, libraries were demultiplexed and amplicons aligned to AMR markers, human reference, and synthetic sequence spike-ins. The number of unique template molecules (estimated deduplicated templates (EDT) detected was estimated from the amplicon alignments for each marker and housekeeping gene and from the unique molecular identifiers for the quality control oligonucleotides. Background EDT levels of each marker and housekeeping gene were estimated per sequencing batch using negative batch controls. An upper bound estimate of background signal was subtracted from the observed EDT for each gene to generate a corrected EDT estimate.

Corrected AMR marker genes and WINC EDT were used as input into the AMR statistical model. For each AMR marker gene, the AMR statistical model identifies all the pathogens detected by the Karius Test known to carry that gene, referred to here as “carriers,” and their respective MPM values. The statistical model then evaluates the likelihood of all the possible combinations of carriers and potential AMR gene copy number given an established prior to output a probability of linkage of AMR to each carrier. For carriers that qualified for the follow-on AMR testing, the Karius Test AMR caller will report: “detected” if there is strong evidence for linkage of AMR presence to the pathogen; “detected-unassigned” if there is strong evidence for AMR presence, but linkage cannot be assigned to one of multiple qualified pathogens; “not detected” if there is strong evidence for linkage of absence of AMR to the pathogen; and “indeterminate” if there is insufficient evidence to support linkage of AMR presence or absence to a qualified pathogen. For *S. aureus*, *S. epidermidis*, and *E. faecium* an additional check of consistency of housekeeping gene EDT to pathogen MPM and AMR EDT is performed. Failure of this check will result in an “indeterminate” report.

Several sample and sequencing batch quality control checks were implemented. EDT of ID Spikes, oligonucleotides unique to each sample, was used to monitor for sample mix-ups and sample cross-contamination. The reaction efficiency of the enrichment process was monitored by the WINC EDT - below 100 WINC EDT, the sample was requeued, unless there was an AMR detection or if an AMR “absence” call is supported by an internal check using EDT of housekeeping genes for *S. aureus, S. epidermidis*, or *E. faecium*. Failure of these sample internal quality control checks leads to an “indeterminate” report for the affected sample and pathogen. Sequencing batch quality control checks included a check that the correct AMR genes were detected/not detected in the positive control samples and that the background EDT for the genes present in the batch controls did not exceed the upper bound of expectations established for the reagent lot. Failure of the sequencing batch quality control checks leads to requeues and “indeterminate” AMR reports for all the affected genes and/or pathogens in the sequencing batch.

### Limits of detection and precision. SCC*mec*

The limit of detection (LoD) of the MRSA/MSSA calls made by the SCC*mec* caller was established by measuring sensitivity using *in-silico* modeling based on an established dilution series of a *S. aureus* genome spiked into a human cfDNA background representing the 10th percentile of human cfDNA concentration observed in commercial samples (11). The 10 half-log dilutions ranged from 316,000 MPM down to 10 MPM, in addition to un-spiked samples. Ten genome assemblies were chosen (5 MRSA and 5 MSSA) that were geographically and genotypically diverse, publicly available, and tested for phenotypic susceptibility (Supplemental Table 1). For each such genome, we created an *in-silico* dilution series from the established dilution series by swapping existing reads with simulated reads from the genome. Using these simulated data, we performed a probit analysis to establish a 95% LoD for each genome after subsampling to the unique WINC QC minimum (25,000) as well as the typical level (300,000) observed in our production samples. The final LoD values presented in *S. aureus* MPM and EDR were averaged across the genomes.

For precision assessment, replicates of clinical samples with a MRSA or MSSA determination by the SCC*mec* caller were reprocessed in the same sequencing batch for repeatability or across sequencing batches for reproducibility (Supplemental Methods).

### AMR gene panel

The LoD for calling AMR presence and absence was calculated using contrived laboratory samples containing multiplexed mixtures of sheared microbial genomes carrying the target AMR genes from the panel (Supplemental Table 2). The multiplexed mixtures were spiked in 8 half-log dilutions into a low human cfDNA plasma background (Supplemental Table 3). The *vanB* variant carried by the ATCC strain used in the genome mix shares only 95% nucleotide identity with the *vanB* used to design targeting primers; 9 of 16 primer-binding sites contain a mutation. To compensate for the loss of signal due to imperfect primer binding sites, *vanB* concentration in the genome mixes was selectively increased five-fold. To establish the LoD for calling AMR(-) genomes, e.g. absence of *mec*A, samples from the dilution series of a complementary genome not carrying the target were used, e.g. for *mecA* absence LoD, *S. aureus mecC*(+) genome was used and for *mecC* absence LoD, *S. aureus mecA*(+) was used. Replicates were tested over 10 assay runs across 10 days to incorporate variance in typical laboratory conditions. The LoD was estimated using a probit analysis and reported in units of pathogen MPM.

To examine how sequencing depth affects the LoD, each read set was subsampled to 300,000, 3.4 million, and 7.8 million reads, representing the 10th, 25th, and 50th percentile of typical clinical sample total read counts (Supplemental Fig. 1), and the LoD was recalculated. Lower sequencing depth is expected to increase the LoD. After evaluating all sample quality control metrics, two replicates were removed from all subsequent analyses that use LoD samples due to failure of the cross-contamination quality control metrics.

For precision assessment, repeatability and reproducibility of the AMR assays were assessed with the contrived samples from the dilution series described above (Supplemental Methods).

### Inclusivity and exclusivity. SCC*mec*

To assess inclusivity, 360 *in-silico* samples were simulated by spiking simulated reads from 36 MRSA-labeled genome assemblies at a particular read count into a healthy human plasma matrix subsampled to the unique WINC QC minimum. The read count was drawn from distribution matched to *S. aureus* calls observed in commercial samples run with the Karius Test, and the database was blinded to the assembly from which the reads were simulated. To assess exclusivity, the same procedure was performed for 11 MSSA-labeled genome assemblies, generating 110 *in-silico* samples. The MSSA-labeled genome assemblies included genomes containing a non-*mec* SCC element that lacks a *mec* resistance gene.

### AMR gene panel

To test the robustness of the primers and bioinformatics protocols to sequence diversity, *in-silico* samples were created using available AMR alleles from curated NCBI nucleotide sequences available in the Comprehensive Antibiotic Resistance Database (CARD) (24). To generate an *in-silico sample*, an allele from the desired on-panel AMR gene was chosen from the set of available nucleotide sequences at random. From the LoD experiment, the empirically tested concentration closest to, but not below, twice the LoD for calling the presence of the gene at median sequencing depth was selected.

In the technical replicate sequencing reads, the AMR target amplicons, and their corresponding AMR target primers, which we refer to as *activated* primers, i.e., primers that successfully amplified a template molecule, were identified. The nucleotide sequences of the amplicons were then swapped with nucleotide sequences corresponding to the chosen AMR allele. As a conservative modeling of primer binding dynamics, only amplicons whose activated primers had a perfect match on the allele were swapped; otherwise, they were discarded from the *in-silico* sample. The inclusivity of housekeeping genes was considered (Supplemental Methods) and found that housekeeping genes have high percent identity matches to all the target species assemblies tested. Given the high inclusivity of housekeeping genes for their target species, the read swapping protocol was not carried out for them. For each on-panel AMR gene, 50 such *in-silico* samples were created.

To evaluate robustness of our primers and bioinformatics protocols to cross-reactive sequences, AMR genes and primers were mapped to microbe, fungi, and viral reference genomes from the Karius Test pathogen reference genome database and the human reference genome to generate a list of cross-reactive sequences for each on-panel AMR gene. High identity matches (>95% identity, >99% coverage) to the AMR target genes or alleles of the AMR target genes were excluded to eliminate true AMR genes from the list of cross-reactive sequences.

In addition, genomes of clinical isolates of organisms in Table 1 with presumed absence of the associated AMR gene(s) were queried using NCBI’s Pathogen Detection database. Absence of AMR genes was judged using NCBI’s AMRFinderPlus annotation for the clinical isolate genome. Only complete genomes were queried in order to have high quality sequence data.

To generate a single *in-silico* sample for an AMR gene, an AMR (-) genome from the appropriate target species or a cross-reactive sequence from the above lists was chosen. Then, the empirically tested concentration closest to, but not below, twice the LoD for calling the absence of the AMR gene at median sequencing depth from the LoD experiment was selected. Using relaxed parameters that allow less-than-perfect primer matches, the activated primers were mapped to the chosen AMR (-) genome or cross-reactive sequence to identify putative primer-binding regions and a “read swapping” protocol was performed as described above. This is a conservative approach that does not fully account for the selectivity of our primers in a reaction. 100 such *in-silico* samples were created per AMR gene (50 with AMR-negative genomes and 50 with putative cross-reactive sequences).

### Clinical validation

A total of 115 unique patient plasma samples which had one of the 18 target pathogens detected in the Karius Test, concordant culture results, applicable orthogonal AST results, and adequate residual volume were included in the clinical validation study. The study sites, number of samples with orthogonal data by site, and the data or sample collection protocols that met the inclusion criteria are shown in Table 2. The orthogonal culture results were obtained from a wide variety of sample types including 71 blood, 17 sputum, 12 BAL, 8 wound, 8 tissue/aspirate, and 4 urine samples. The samples contained 12/18 (66.7%) of the target bacteria including 58 *Staphylococcus aureus*, 16 *Pseudomonas aeruginosa*, 14, *Escherichia coli*, 6 *Klebsiella pneumoniae*, 5 *Serratia marcescens*, 5 *Enterobacter cloacae* complex, 4 *Enterococcus faecium*, 2 *S. epidermidis*, 2 *E. faecalis*, 1 *Proteus mirabilis*, 1 *K. oxytoca* and 1 *Salmonella enterica*.

**Table 2.**
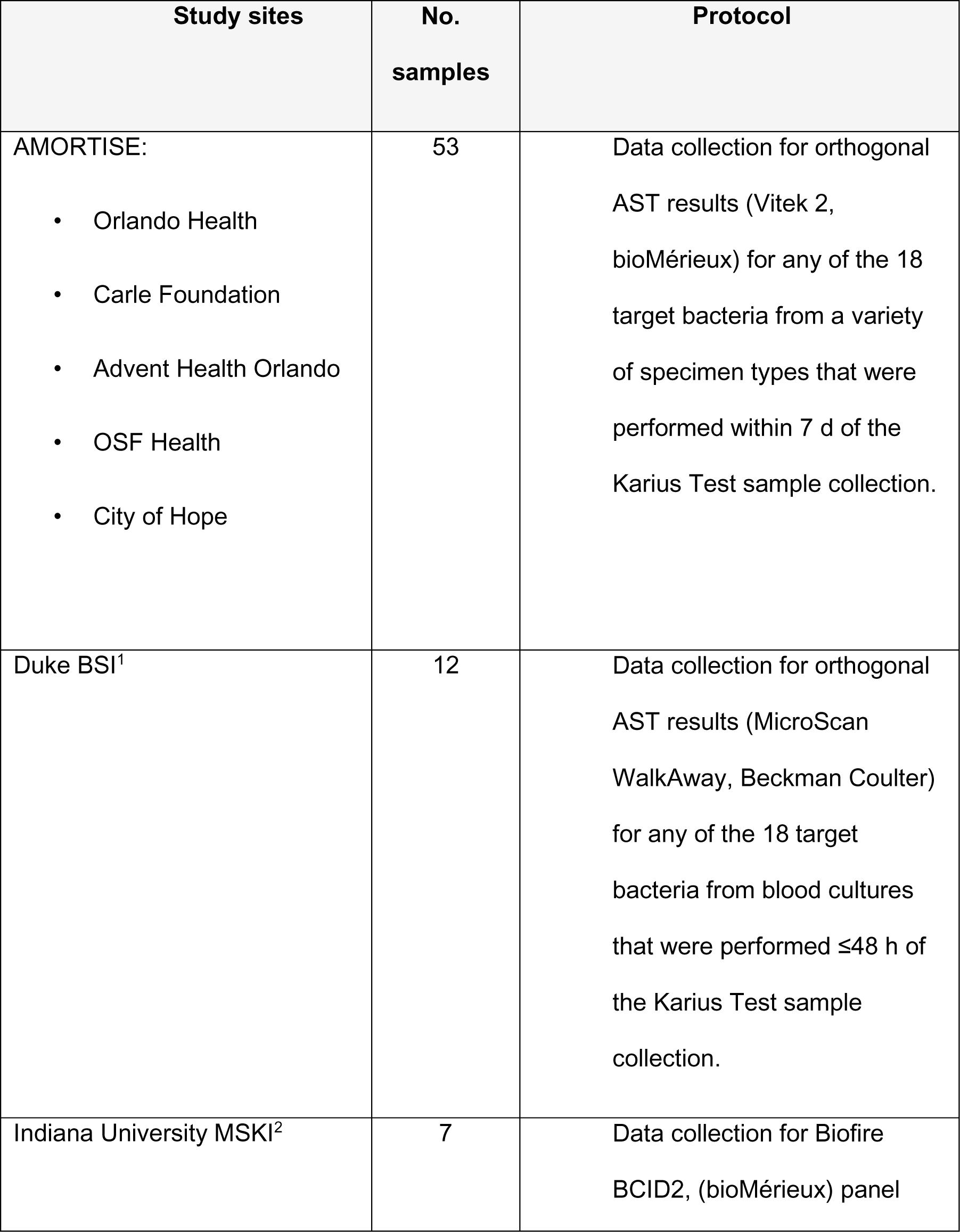
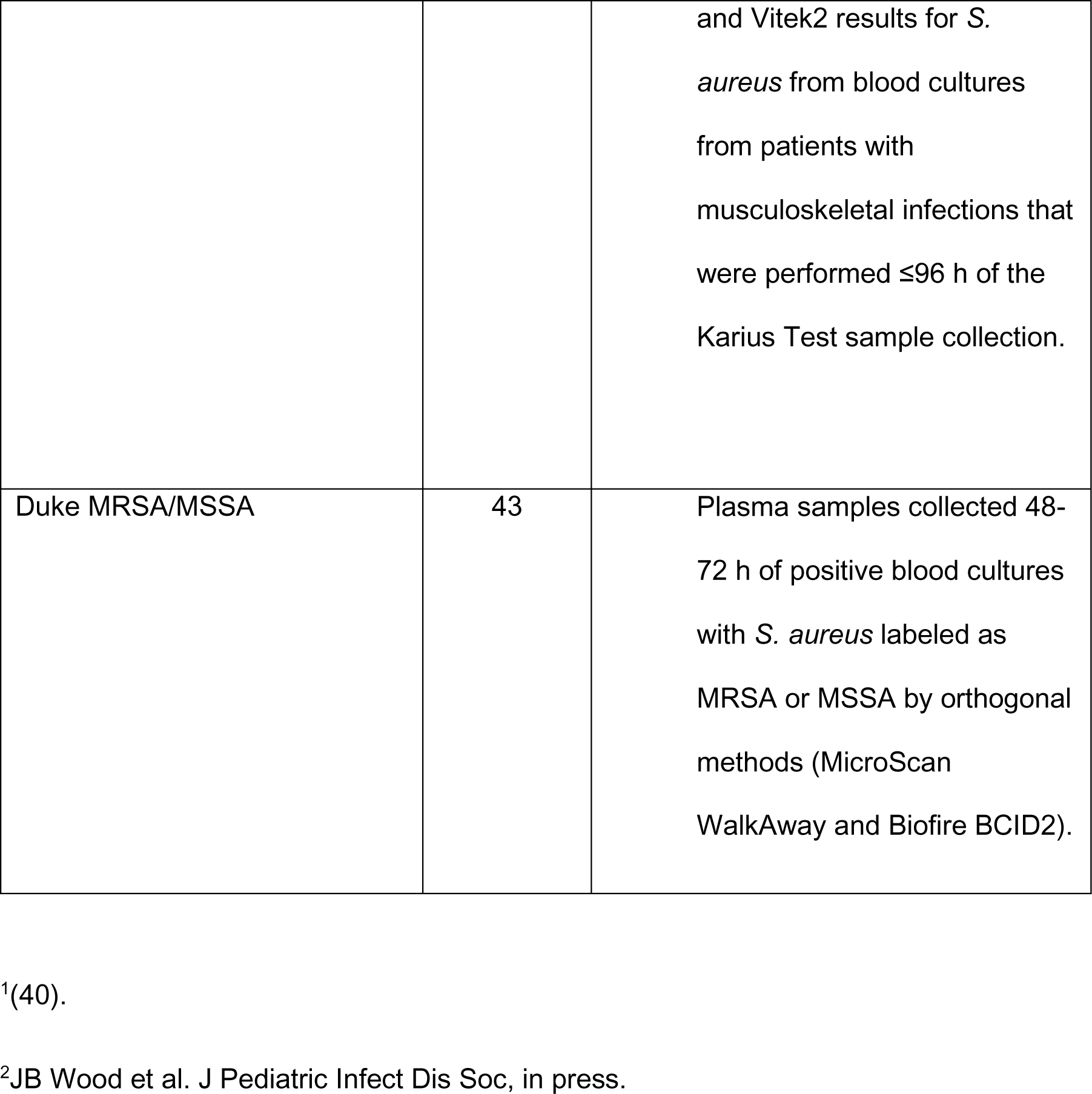
Study sites providing data or specimens for the clinical validation of the Karius Test AMR markers.

The following rules were used to assign labels to the plasma samples included in the analysis: 1) Positive culture for one of the panel staphylococci and reported resistant or susceptible to oxacillin for SCC*mec* and *mecA* or *mecC*. 2) Positive culture for *E. faecalis* or *E. faecium* and reported resistant or susceptible to vancomycin for *vanA* or *vanB*. 3) Positive culture for one of the panel gram-negative bacilli and reported resistant to any one or more of the following antimicrobials tested: cefotaxime, ceftazidime, ceftriaxone, cefepime or aztreonam for putative *bla*_CTX-M_ positives and reported susceptible or intermediate to all key indicator drugs for putative *bla*_CTX-M_ negatives. Not all drugs were tested for each pathogen. 4) Positive culture for one of the panel gram-negative bacilli and reported resistant to any one or more of the carbapenems tested: doripenem, ertapenem, imipenem, or meropenem for putative *bla*_KPC_ positives and reported susceptible to all carbapenems tested for putative *bla*_KPC_ negatives. As above, not all drugs were tested for each pathogen. Overall, for those plasma samples associated with concordant cultures that revealed strains with different AST results, the most resistant phenotype was used as the label.

The orthogonal phenotypic AST was performed by Vitek2 for 52.2% and by MicroScan WalkAway system for 47.8% of samples included in the clinical validation.

Orthogonal genotypic results were also reported for 11 of the AMR On Remnant TISsuE (AMORTISE) Study samples from positive blood cultures including 2 Verigene, 5 GeneMark and 4 Biofire BCID panel results. All the Duke MRSA/MSSA and the Indiana MSKI study samples had associated BCID panel results reported for positive blood cultures for *S. aureus*.

### Statistical Analysis

All statistical analyses were performed in Python 3.10. The LoD was calculated using a maximum-likelihood probit model fit to the data. This model assumed that the number of positives at each concentration followed a binomial distribution and the success rate of the binomial distribution at each concentration was determined by the value of the cumulative normal distribution at that concentration. The parameters of the cumulative normal distribution (probit fit) associating concentration with sensitivity were fitted to maximize the likelihood of the dilution series. The LoD concentration was determined using the probit (inverse of cumulative normal distribution) at 95% sensitivity. The 95% confidence intervals on proportions were calculated using the Clopper-Pearson interval. Descriptive statistics used to assess test performance characteristics, positive percent agreement (PPA), negative percent agreement (NPA), and overall percent agreement (CA) were calculated by accepted methods (25). Diagnostic yield (DY) was defined as the percentage of tests that yielded an actionable result either detected or not detected.

### Ethical considerations

The study was conducted in compliance with good clinical practice guidelines set forth by the International Conference on Harmonization (ICH-E6). The following Institutional Review Boards granted waiver of consent for the collection of additional clinical data to support this study: AdventHealth Institutional Review Board, City of Hope Institutional Review Board, Orlando Health Institutional Review Board, Peoria Institutional Review Board, and Advarra Institutional Review Board. The Indiana University Institutional Review Board and the Duke Institutional Review Board granted study approval with the ability to use the study samples for additional future research upon consent. In addition, the sample identifiers reported here we unknown to anyone outside of the research group.

## RESULTS

### Limit of detection and precision. SCC*mec*

To estimate LoD, multiple genome assemblies and two levels of sequencing depths, representing the minimum for absolute quantification and typical sequencing depth, were simulated in an *in-silico* dilution derived from a representative dilution series of a *S. aureus* sheared genome in a human plasma background. An *in-silico* approach allowed for incorporation of multiple variables, e.g., different MRSA and MSSA genomes and sequencing depths, that the original dilution series did not address. The LoD values at different sequencing depths and SCC*mec* presence or absence were averaged across the genome assemblies assessed (Table 3). Sequencing depth was measured using the number of unique WINC molecules in the sample as a proxy, with the minimum depth 10-fold lower than the typical depth. The presence and absence LoD values were sensitive to sequencing depth, with the LoD increasing approximately 10-fold at the minimum depth. This is expected, as higher pathogen abundance is needed to make a determination in a shotgun sequenced sample with fewer reads. In a typical production sample, the presence LoD is 3,916 MPM and absence LoD is 683 MPM. 100% repeatability and reproducibility were observed in the MRSA and MSSA clinical samples (n=6 per genotype).

**Table 3.** Limit of detection characterization for SCC*mec*.

|  | Average Limit of Detection |  |
| --- | --- | --- |
| Sequencing depth | *SCCmec* presence (MRSA) | *SCCmec* absence (MSSA) |
| QC minimum<br>(25,000 unique WINC) | 54,803 MPM / 4,567 EDR<sup>1</sup> | 10,304 MPM / 858 EDR |
| Standard<br>(300,000 unique WINC) | 3,916 MPM / 3,916 EDR | 683 MPM / 683 EDR |

<sup>1</sup>Estimated deduplicated reads (EDR), represent the number of unique DNA sequencing reads from the reported microorganism present in the sequenced library (relative quantity).

### AMR gene panel

To estimate LoD for each of the targeted AMR genes, a dilution series was created using sheared microbial genomes carrying AMR genes spiked into human plasma background. This dilution series was computationally downsampled to assess LoD at different sequencing depths. Table 4 shows the LoD based on probit model fits for non-downsampled data, and then the data downsampled to each of the predefined sequencing depths. Overall, the presence LoDs range from 425-6,107 MPM and the absence LoDs range from 472-22,680 MPM. LoD was not strongly sensitive to lower sequencing depths - this is because the targeted amplification method creates many amplicons from the original fragment making “surplus” signal. Presence LoD is largely influenced by the gene copy number and gene variant. The *vanB* presence LoD is a conservative estimate, a limitation imposed by the *vanB* gene variant in the genome mixture (see Methods). Since only one microbial genome was assessed to determine the gene LoD, LoD was also characterized assuming one copy of the AMR gene (Supplemental Table 4), to give a conservative upper bound for the LoD as several of the panel AMR genes were carried in multiple copies by the microbes (Supplemental Table 2). For AMR genes that were carried in multiple copies, except *bla*_CTX-M_, LoD increased when assuming one copy. Decrease in *bla*_CTX-M_ LoD at one copy number is due to elimination of positive calls at low pathogen MPM due to reagent background (Supplemental Fig. 2) once the *bla*_CTX-M_ signal was scaled to simulate one gene copy, leading to a different probit model fit (Supplemental Fig. 3).

**Table 4.** LoD by target and sequencing depth for the AMR gene panel.

| Target | Original sequencing depth | 50th percentile sequencing depth | 25th percentile sequencing depth | 10th percentile sequencing depth |
| --- | --- | --- | --- | --- |
| *mecA* presence | 662 MPM | 674 MPM | 966 MPM | 1,712 MPM |
| *mecA* absence | 2,316 MPM | 2,316 MPM | 2,177 MPM | 6,694 MPM |
| *mecC* presence | 808 MPM | 856 MPM | 1,180 MPM | 1,803 MPM |
| *mecC* absence | 472 MPM | 472 MPM | 472 MPM | 1,000 MPM |
| *vanA* presence | 425 MPM | 372 MPM | 437 MPM | 646 MPM |
| *vanA* absence | 1,044 MPM | 1,044 MPM | 2,617 MPM | 1,000 MPM |
| *vanB* presence* | 4,985 MPM | 4,996 MPM | 5,000 MPM | 6,730 MPM |
| *vanB* absence | 1,074 MPM | 1,105 MPM | 1,105 MPM | 1,180 MPM |
| *bla*<sub>CTX-M</sub> presence | 6,107 MPM | 6,633 MPM | 5,793 MPM | 6,971 MPM |
| *bla*<sub>CTX-M</sub> absence | 22,680 MPM | 19,640 MPM | – | 3,471 MPM |
| *bla*<sub>KPC</sub> presence | 1,346 MPM | 1,346 MPM | 1,380 MPM | 1,769 MPM |
| *bla*<sub>KPC</sub> absence | 1,555 MPM | 1,555 MPM | 1,555 MPM | 4,672 MPM |

For *bla*_CTX-M_, the empirically measured LoD was also impacted by the presence of reagent background signal (Supplemental Fig. 2), especially the absence LoD. For *bla*_CTX-M_, downsampling to lower sequencing depths decreased the *bla*_CTX-M_ reagent background signal, leading to more effective background subtraction and improved probit fits to the observed data. *bla*_CTX-M_ absence LoD was unable to be estimated using a probit model for samples downsampled to the 25^th^ percentile of sequencing depth because there was no concentration at which there was a ≥95% absence call rate.

100% repeatability and reproducibility were observed when evaluating precision in the contrived samples (n=10 to 20 per target).

### Inclusivity and exclusivity

Different alleles of AMR genes may exhibit different sensitivities in the AMR assay due to primer-template mismatches or failure to successfully align to the reference sequences in the bioinformatics pipeline. To assess a wide array of alleles representative of the range of variation present in the population, *in-silico* simulations were performed, derived from the contrived samples of the LoD dilution series. Across the different alleles assessed, 100% PPA was observed for all targets except *bla*_CTX-M_ (Table 5). Inclusivity for *bla*_CTX-M_ was limited to 54% in this analysis (based on a single sampling of 50 alleles) because the targeted approach has primers with perfect homology only to a single clade of *bla*_CTX-M_ variants (*bla*_CTX-M-3_ clade), but not the other highly divergent clades (Fig. 2). In general, based on the prevalences derived from CARD resistomes dataset and literature, the sensitivity for *bla*_CTX-M_ variants is expected to be 67-75% (26). Common *bla*_CTX-M_ variants captured by our AMR gene enrichment include *bla*_CTX-M-3_, *bla*_CTX-M-15_, *bla*_CTX-M-32_, *bla*_CTX-M-55_, and *bla*_CTX-M-1_. Common CTX-M variants poorly captured or missed by AMR gene enrichment include *bla*_CTX-M-2_, *bla*_CTX-M-9_, *bla*_CTX-M-14_, *bla*_CTX-M-27_, and *bla*_CTX-M-65_. Except for the highly divergent *bla*_CTX-M_, the AMR assay is largely insensitive to variation in gene sequences.

**Table 5.** Inclusivity by AMR marker.

| Target | Inclusivity | 95% CI |
| --- | --- | --- |
| *SCC<sub>mec</sub>* | 100% (36/36) | 90%-100% |
| *mecA* | 100% (50/50) | 93%-100% |
| *mecC* | 100% (50/50) | 93%-100% |
| *vanA* | 100% (50/50) | 93%-100% |
| *vanB* | 100% (50/50) | 93%-100% |
| *bla<sub>CTX-M</sub>* | 54% (27/50) | 39%-68% |
| *bla<sub>KPC</sub>* | 100% (50/50) | 93%-100% |

**Fig. 2.**
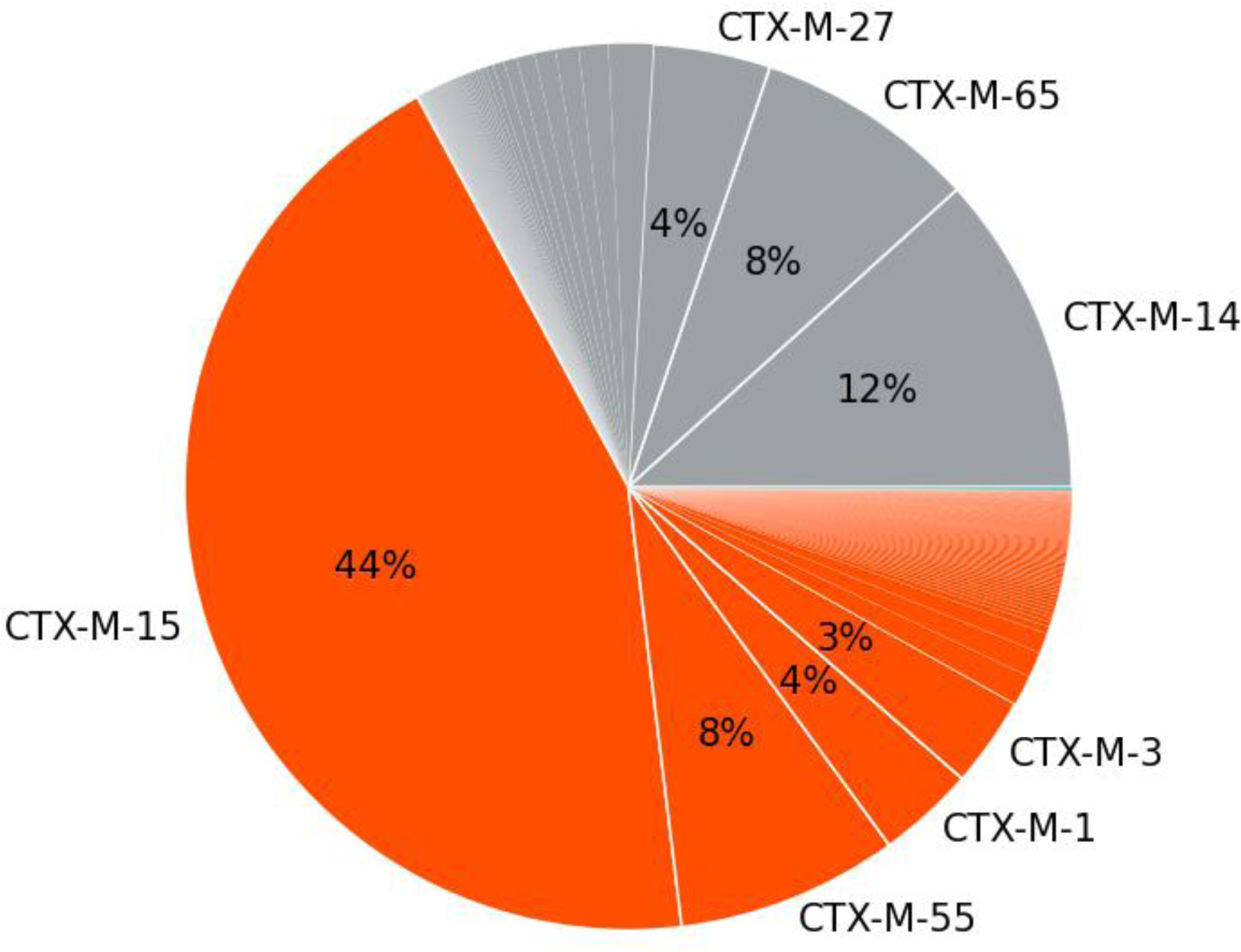
*bla*_CTX-M_ variant prevalence (%) taken from CARD Resistomes database. Orange represents CTX-M variants with >95% sequence identity to the reference CTX-M sequence used in primer design and are captured well by the AMR assay primers. Gray represents CTX-M variants with 70-80% sequence identity that are not captured by the AMR assay primers.

The ability of the Karius Test AMR detection assays to call absence may be impeded by the presence of non-AMR sequences that are a match for the primers and similar in sequence to the AMR gene. Like the inclusivity assessment above, an *in-silico* approach was chosen to assess a wide array of possible cross-reactive sequences. An exclusivity of 100% was observed (Table 6), showing a remarkably high assay specificity for the AMR markers tested. From a literature and database search, we identified allotypes of *mecC*, *mecC1* and *mecC2* (27, 28), which have 93.7% and 96.3% sequence identity to *mecC*, respectively. When simulating samples using these allotypes, *mecC* was called in all simulations, due to the extremely high sequence similarity. Only *mecC1* is an allotype known to be phenotypically susceptible to methicillin (27). This allotype has the potential to affect exclusivity, but it is expected to be extremely rare in the patient population, given that *mecC* itself has a low prevalence in patients (0-3%) (29). Overall, exclusivity is highly unlikely to be impacted by any cross-reactive sequence with <95% sequence identity to the AMR markers.

**Table 6.** Exclusivity by AMR marker.

| Target | Exclusivity | 95% CI |
| --- | --- | --- |
| SCC<sub>mec</sub> | 100% (11/11) | 71%-100% |
| *mecA* | 100% (100/100) | 96%-100% |
| *mecC* | 100% (100/100) | 96%-100% |
| *vanA* | 100% (100/100) | 96%-100% |
| *vanB* | 100% (100/100) | 96%-100% |
| *bla*<sub>CTX-M</sub> | 100% (100/100) | 96%-100% |
| *bla*<sub>KPC</sub> | 100% (100/100) | 96%-100% |

### Clinical validation

The positive percent agreement (PPA), negative percent agreement (NPA), overall percent agreement (OPA), and diagnostic yield (DY) and 95% confidence intervals in brackets were estimated for each maker. All data for clinical validation samples are shown in Supplemental Table 7.

Correlation of SCC*mec* results with phenotype for 58 *S. aureus* detections are shown in Table 7. The PPA, NPA, OPA and DY were 13/13 (100%) [73.5-100%], 17/18 (95.0%) [72.7-99.8%], 30/31 (96.8%) [83.3-99.9%] and 31/58 (53.4%) [39.8-66.6%], respectively. In the one false positive both SCC*mec* and *mecA* were detected and likely represents a true positive. The single false negative remained a false negative after review.

**Table 7.** Correlation of SCC*mec* results with phenotype for 58 *Staphylococcus aureus* detections. The 95% confidence intervals for performance metric estimates are in brackets.

| SCC*mec* result | Phenotype |  |
| --- | --- | --- |
|  | Methicillin resistant | Methicillin susceptible |
| Detected | 13 | 1<sup>a</sup> |
| Not detected | 0 | 17 |
| Indeterminate | 12 | 15 |
| Agreement | PPA: 13/13 (100%)<br>[73.5-100%] | NPA: 17/18 (94.4%)<br>[72.7-99.8%] |
| Overall Agreement | 30/31 (96.8%) [83.3-99.9%] |  |
| Diagnostic Yield | 31/58 (53.4%) [39.8-66.6%] |  |

<sup>a</sup>Both SCC*mec* and *mecA* were detected.

Correlation of *mecA* results with phenotype for the same 58 *S. aureus* detections are shown in Table 8. The PPA, NPA, OPA, and DY were 14/15 (93.3%) [68.0-99.8%], 19/20 (95.0%) [75.1-99.8%], 33/35 (94.3%) [80.8-99.3%] and 35/58 (60.3%) [46.6-72.9%], respectively. The single false positive and false negative occurred with the same two strains described above for SCC*mec*. No *mecC*-positive strains were detected.

**Table 8.**
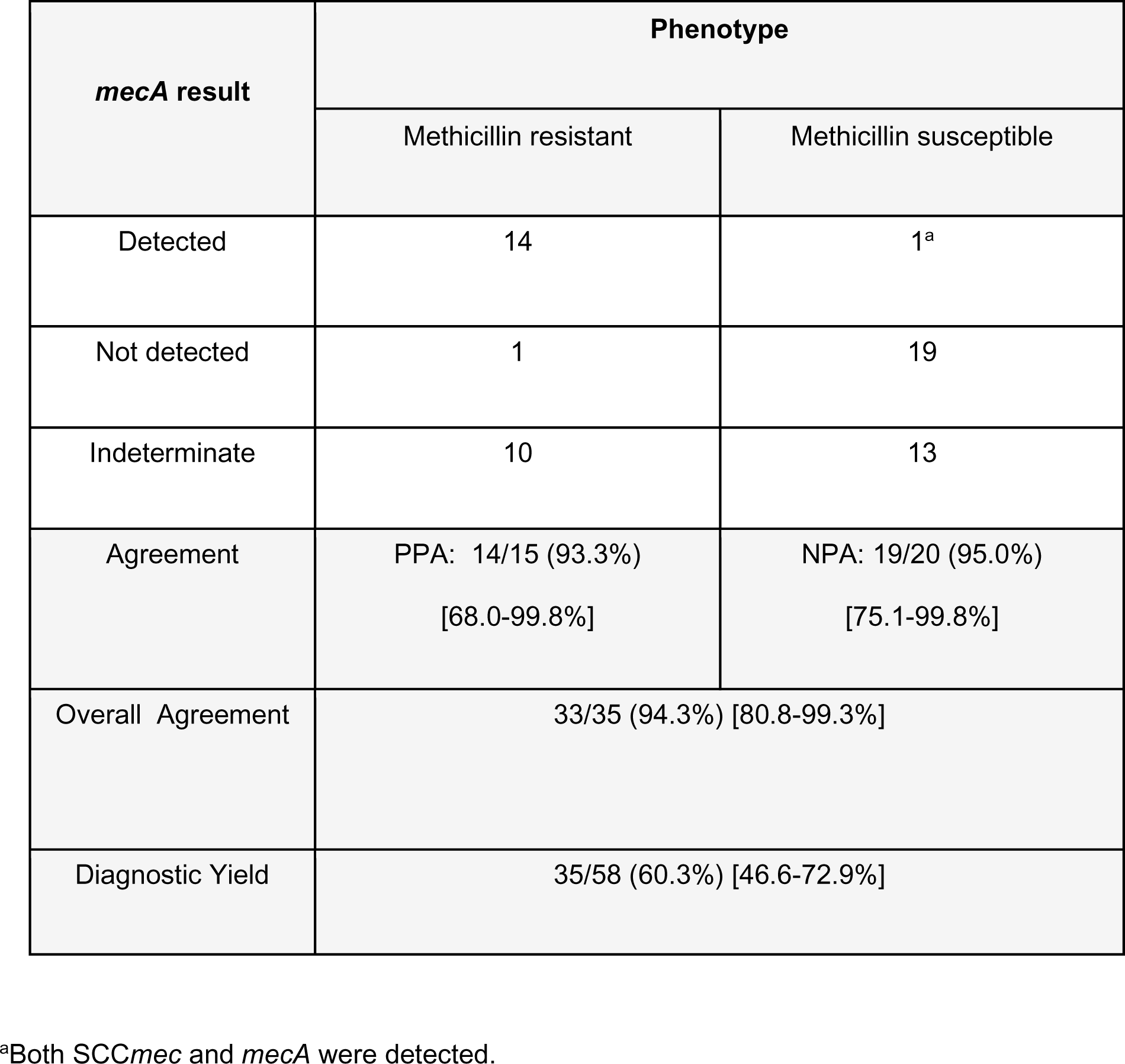
Correlation of *mecA* results with phenotype for 58 *Staphylococcus aureus* detections. The 95% confidence intervals for performance metric estimates are in brackets.

The results when the SCC*mec* and *mecA* tests were combined for 58 *Staphylococcus aureus* and 2 *S. epidermidis* detections in the decision workflow for methicillin resistance in staphylococci are shown in Table 9. As expected, the PPA, NPA and CA were nearly identical to what was observed when each target was assessed separately. However, the DY increased to 70% [56.8-81.1%].

**Table 9.**
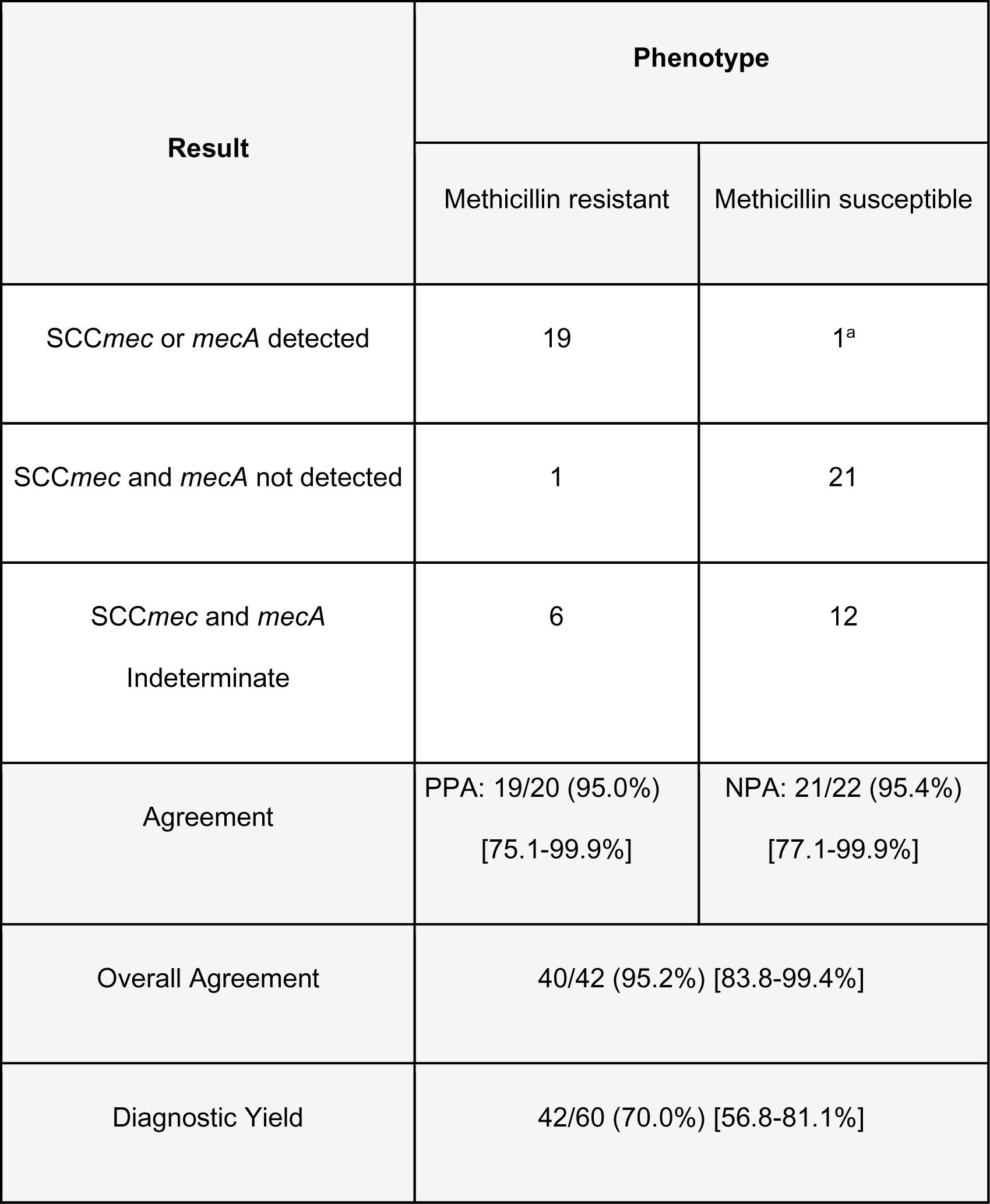
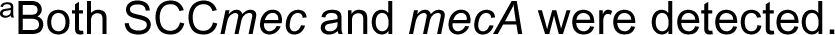
Correlation or results when SCC*mec* and *mecA* tests are combined for 58 *Staphylococcus aureus* and 2 *S. epidermidis* detections in the decision workflow for methicillin resistance in staphylococci. The 95% confidence intervals for performance metric estimates are in brackets.

| Result | Phenotype |  |
| --- | --- | --- |
|  | Methicillin resistant | Methicillin susceptible |
| SCC*mec* or *mecA* detected | 19 | 1<sup>a</sup> |
| SCC*mec* and *mecA* not detected | 1 | 21 |
| SCC*mec* and *mecA*<br>Indeterminate | 6 | 12 |
| Agreement | PPA: 19/20 (95.0%)<br>[75.1-99.9%] | NPA: 21/22 (95.4%)<br>[77.1-99.9%] |
| Overall Agreement | 40/42 (95.2%) [83.8-99.4%] |  |
| Diagnostic Yield | 42/60 (70.0%) [56.8-81.1%] |  |

<sup>a</sup>Both *SCCmec* and *mecA* were detected.

Correlation of *vanA* results with phenotype for 6 *Enterococcus spp.* detections (4 *E. faecium* and 2 *E. faecalis*) are shown in Table 10. The PPA, NPA, OPA, and DY were 3/3 (100%) [29.2-100%], 2/2 (100%) [15.8-100%], 5/5 (100%) [47.8-100%] and 5/6 (83.3%) [35.8-99.5%], respectively. No *vanB*-positive strains were detected.

**Table 10.**
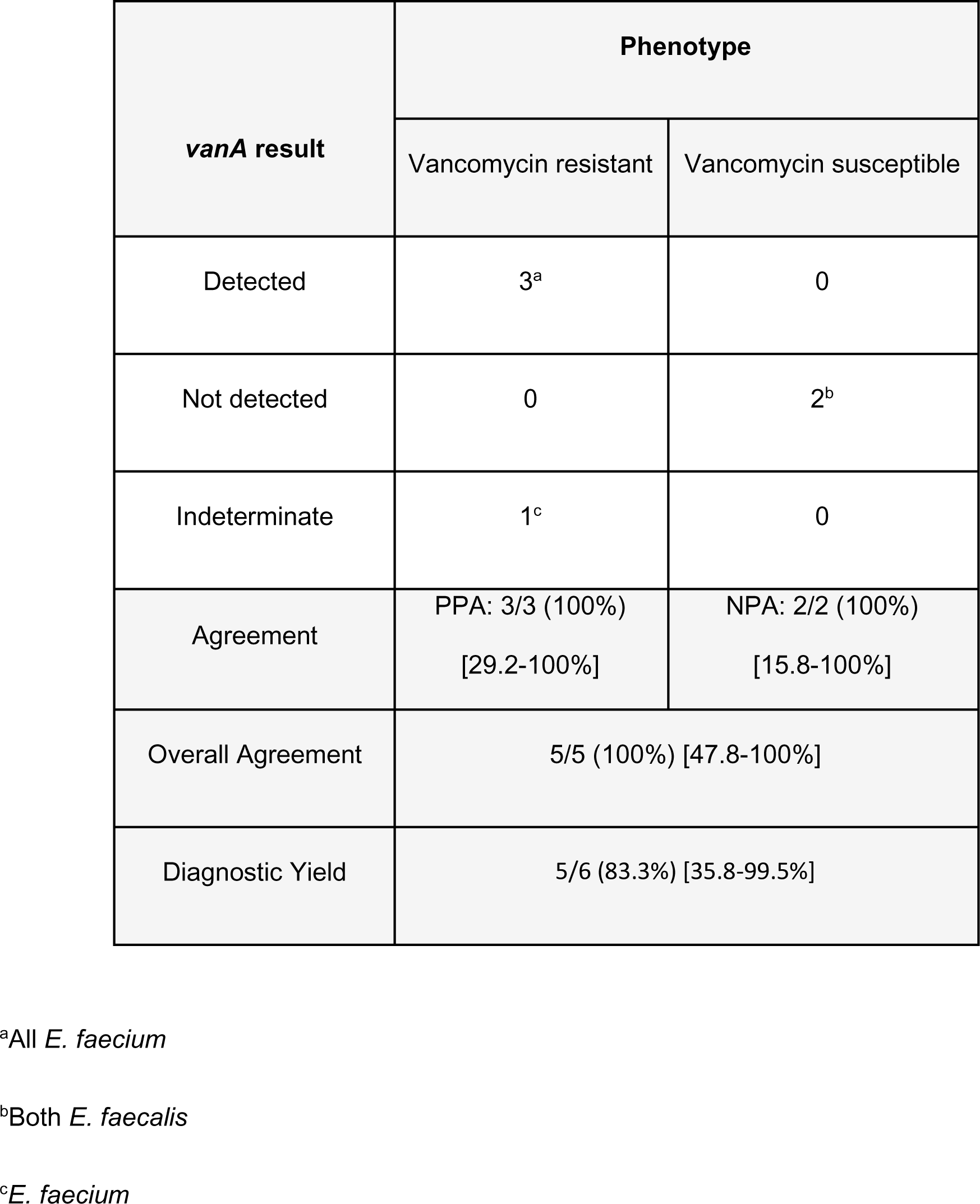
Correlation of *vanA* results with phenotype for 6 *Enterococcus spp.* detections. The 95% confidence intervals for performance metric estimates are in brackets.

Correlation of *bla*_CTX-M_ results with ESBL phenotype for 49 Gram-negative bacilli detections are shown in Table 11. The PPA, NPA, OPA and DY were 5/9 (55.5%) [21.2-86.3%], 26/26 (100%) [86.7-100%], 31/35 (88.5%) [73.2-96.8%] and 35/49 (71.4%) [56.7-83.4%], respectively. The 5 true positive *bla*_CTX-M_ detections included 4 *E.coli* and 1 *K.pneumoniae*.

**Table 11.**
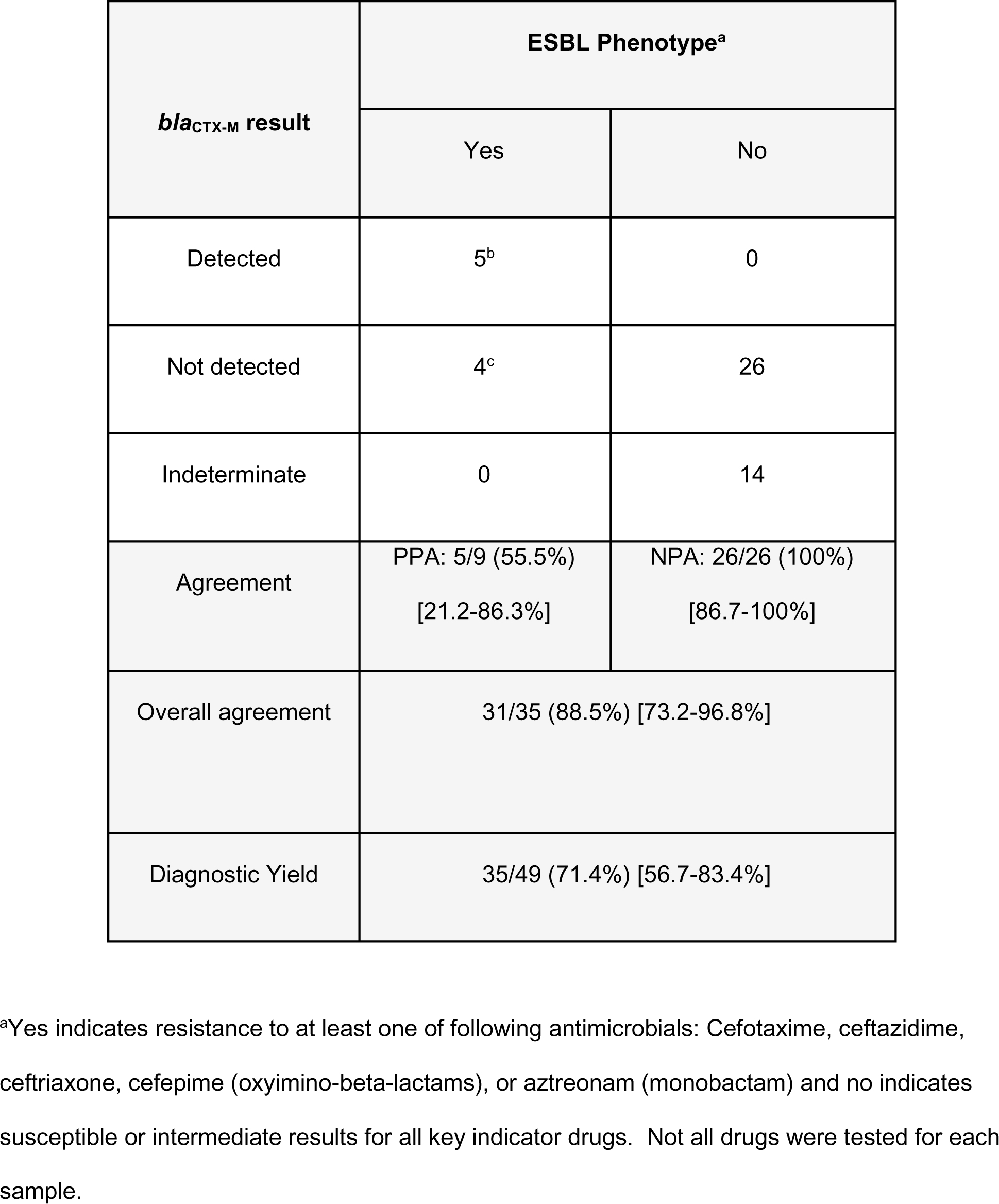
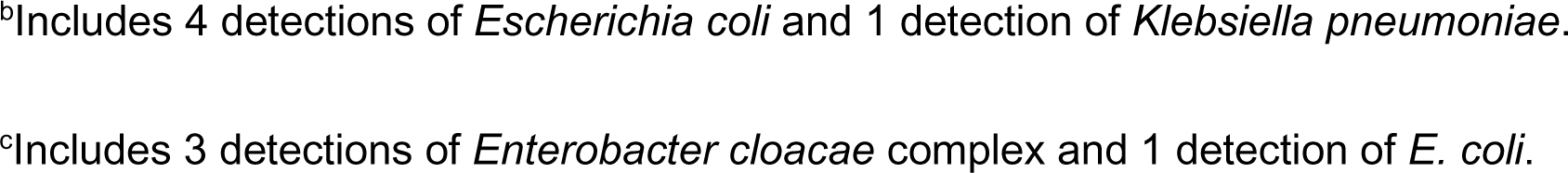
Correlation of *bla*_CTX-M_ results with ESBL phenotype for 49 Gram-negative bacilli detections. The 95% confidence intervals for performance metric estimates are in brackets.

| *bla*<sub>CTX-M</sub> result | ESBL Phenotype<sup>a</sup> |  |
| --- | --- | --- |
|  | Yes | No |
| Detected | 5<sup>b</sup> | 0 |
| Not detected | 4<sup>c</sup> | 26 |
| Indeterminate | 0 | 14 |
| Agreement | PPA: 5/9 (55.5%)<br>[21.2-86.3%] | NPA: 26/26 (100%)<br>[86.7-100%] |
| Overall agreement | 31/35 (88.5%) [73.2-96.8%] |  |
| Diagnostic Yield | 35/49 (71.4%) [56.7-83.4%] |  |

<sup>a</sup>Yes indicates resistance to at least one of following antimicrobials: Cefotaxime, ceftazidime, ceftriaxone, cefepime (oxymino-beta-lactams), or aztreonam (monobactam) and no indicates susceptible or intermediate results for all key indicator drugs. Not all drugs were tested for each sample.
<sup>b</sup>Includes 4 detections of *Escherichia coli* and 1 detection of *Klebsiella pneumoniae*.
<sup>c</sup>Includes 3 detections of *Enterobacter cloacae* complex and 1 detection of *E. coli*.

The 4 false negative *bla*_CTX-M_ detections included 3 *E. cloacae* complex detections and 1 detection of *E. coli*. ESBLs are not readily distinguishable phenotypically from each other and de-repressed or plasmid-encoded AmpC β*-*lactamase*s*. Susceptibility to cephamycins can help distinguish between them but data were unavailable for these strains. However, ESBL and AmpC β-lactamases can be distinguished phenotypically based on differential susceptibility to cefepime. Cefepime is usually not hydrolyzed by AmpC β-lactamases whereas ESBL producers most often have elevated cefepime MICs and are interpreted as susceptible dose-dependent or resistant to cefepime (30).

Two of the three *E. cloacae* complex and the single *E. coli* strains were susceptible to cefepime. The remaining *E. cloacae* complex detection was resistant to cefepime, ceftriaxone and aztreonam. The resolution of the four false negatives using these revised criteria is shown in Table 12. Following resolution of false negative result discrepancies, the PPA of *bla*_CTX-M_ increased from 55.5% to 83.3% and the OPA increased from 88.5% to 97.1%. The other performance metrics remained the same. As a check on the revised criteria for labeling samples we observed that all five true positive *bla*_CTX-M_ detections were from strains that were resistant to cefepime.

**Table 12.**
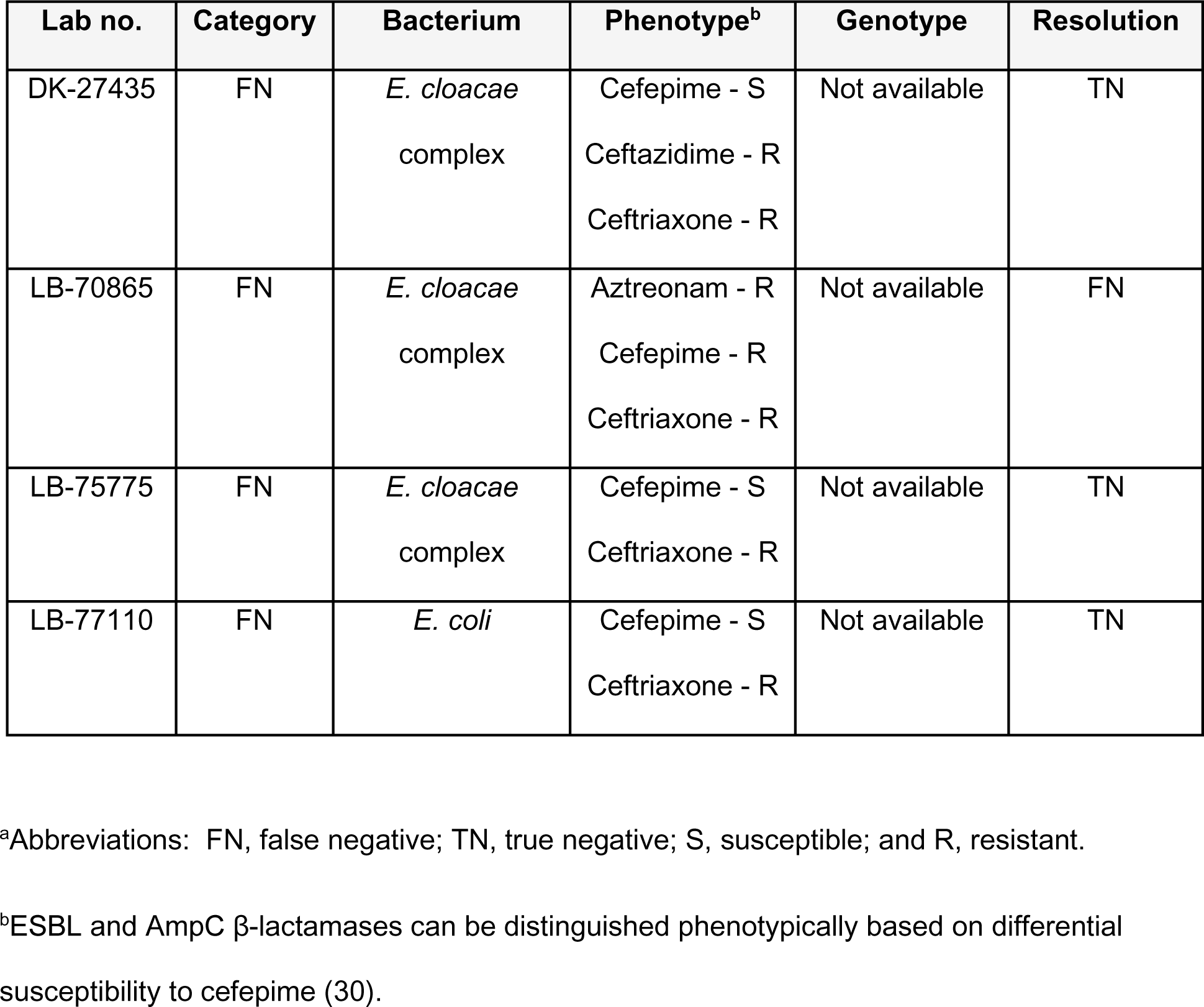
Resolution of *bla*_CTX-M_ false negative detections^a^.

Correlation of *bla*_KPC_ results with carbapenemase phenotype for 44 Gram-negative bacilli detections are shown in Table 13. The PPA, NPA, OPA, and DY were 0/2 (0%) [0-84.2%], 23/23 (100%) [85.1-100%], 23/25 (92.3%) [73.9-99.0%] and 25/44 (56.8%) [41.0-71.6%], respectively. There were no true positive results. The 2 false negative results included 1 detection of *P. aeruginosa* by the Karius Test in a patient who had two cultures positive for strains with different carbapenem phenotypes (BAL meropenem susceptible and sputum meropenem resistant) and one *E. coli* detection that was that was resistant to both ertapenem and meropenem. The most resistant *P. aeruginosa* strain was selected to label the specimen, but the strain from the BAL was deemed more likely to be associated with disease and therefore most likely a true negative. The false negative detection for the *E. coli* could not be resolved.

**Table 13.**
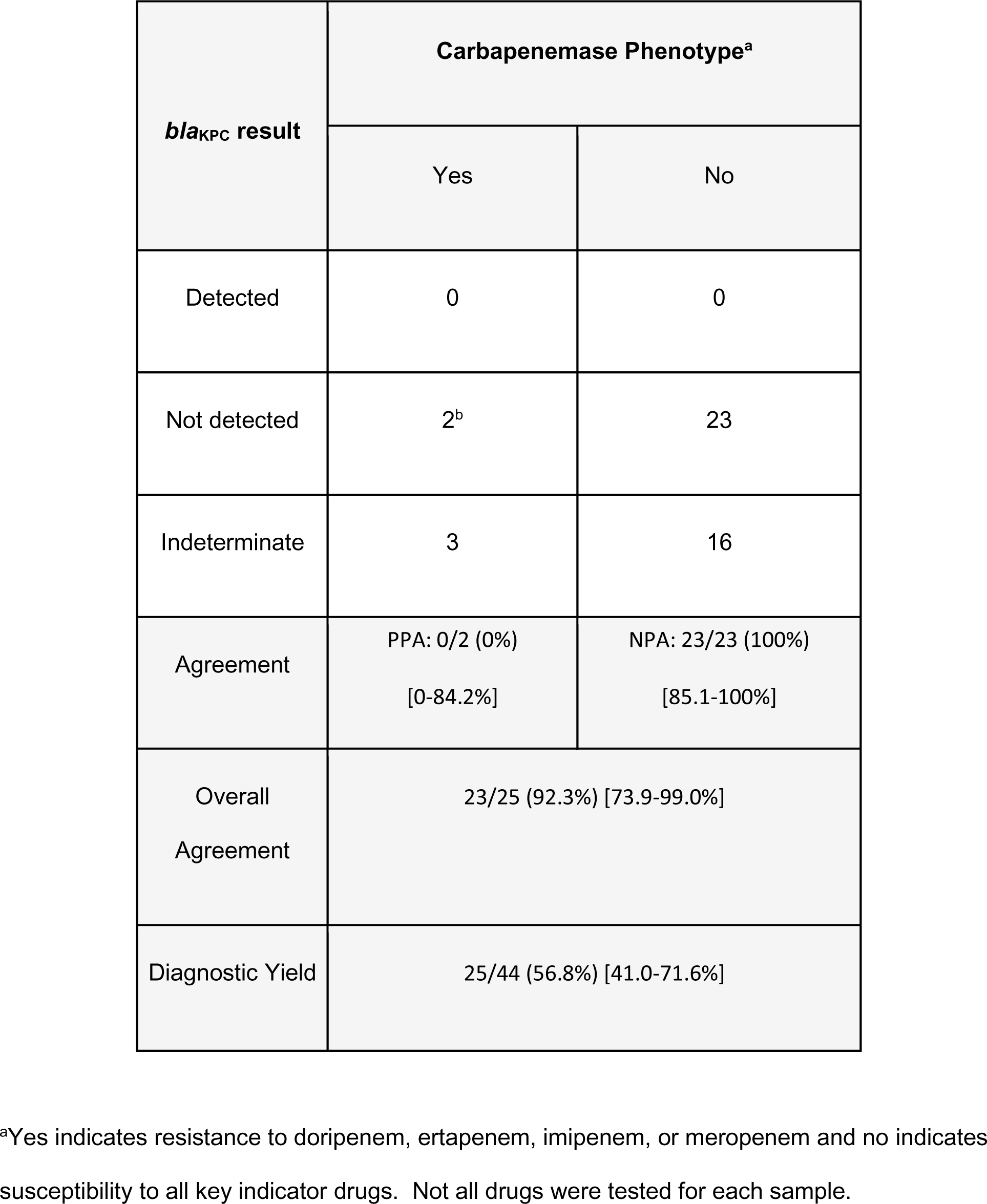
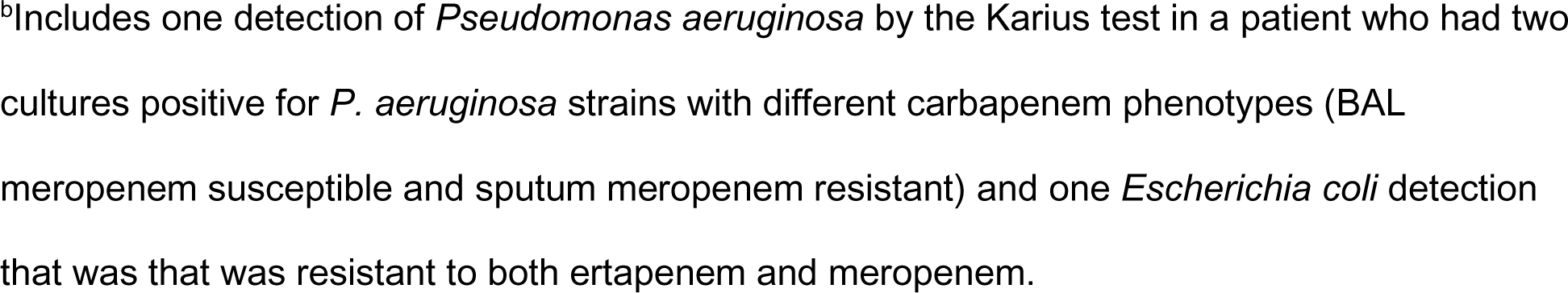
Correlation of *bla*_KPC_ results with carbapenemase phenotype for 44 Gram-negative bacilli detections. The 95% confidence intervals for performance metric estimates are in brackets.

| *bla*<sub>KPC</sub> result | Carbapenemase Phenotype<sup>a</sup> |  |
| --- | --- | --- |
|  | Yes | No |
| Detected | 0 | 0 |
| Not detected | 2<sup>b</sup> | 23 |
| Indeterminate | 3 | 16 |
| Agreement | PPA: 0/2 (0%)<br>[0-84.2%] | NPA: 23/23 (100%)<br>[85.1-100%] |
| Overall Agreement | 23/25 (92.3%) [73.9-99.0%] |  |
| Diagnostic Yield | 25/44 (56.8%) [41.0-71.6%] |  |

<sup>a</sup>Yes indicates resistance to doripenem, ertapenem, imipenem, or meropenem and no indicates susceptibility to all key indicator drugs. Not all drugs were tested for each sample.
<sup>b</sup>Includes one detection of *Pseudomonas aeruginosa* by the Karius test in a patient who had two cultures positive for *P. aeruginosa* strains with different carbapenem phenotypes (BAL meropenem susceptible and sputum meropenem resistant) and one *Escherichia coli* detection that was resistant to both ertapenem and meropenem.

## DISCUSSION

Clinical metagenomic sequencing is rapidly evolving and has the potential for detection of microbial genomes directly in clinical samples. Moreover, sequencing of mcfDNA from plasma has the distinct advantage of not requiring samples of infected body fluids or tissues which often require invasive procedures to collect. There are currently no such FDA-cleared assays for detection of bacterial pathogens and resistance genes/determinants; however, this field is expected to advance quickly, given the rapid advances in technology and the clinical need. This is the first report on the use of plasma mcfDNA sequencing to identify AMR genetic markers for prediction of AMR.

The rarity and short fragment lengths of mcfDNA in plasma (31) present significant technical challenges for detection of AMR markers. Another challenge is the small size of the AMR markers in relation to bacterial genome sizes. The SCC*mec* marker ranges in size from 21 to 60 kb (23) and the individual AMR target genes range from approximately 900 to 2000 bp (24). While the analytical and clinical accuracy of the SCC*mec* detection was extremely high, the diagnostic yield was relatively low, at approximately 50%, because there is no enrichment for SCC*mec*. Target amplification increased the diagnostic yield of the *mecA* gene detection and added the ability to call additional AMR markers in mcfDNA. Other NGS targeting methods starting with isolates, cultures, or whole genomes have also been described (5, 32, 33, 35).

Despite substantial technical challenges we were able to develop and incorporate robust assays for AMR markers into the Karius Test. The LoDs for both the presence and absence of all AMR markers were determined. In a typical production sample (unique WINC 300,000), the presence LoD is 3,916 MPM and absence LoD is 683 MPM for the SCC*mec*. The higher presence LoD is likely due to interfering species in the human background which cause no-calls for MRSA assemblies when SCC*mec* and *S. aureus* are low. In addition, the probit fits did not fit the observed data patterns very well for MSSA because there was a sharp boundary between no absence calls and >95% absence calls.

Overall, the presence LoDs ranged from 425 to 6,107 MPM and the absence LoDs ranged from 472 to 22,680 MPM for the single AMR gene assays. The *vanB* and *bla*_CTX_ assays represented the biggest LoD challenges for the presence and absence of these genes at 6,107 and 22,680 MPM, respectively. The *vanB* variant carried by the ATCC strain used in the LoD genome mix is an imperfect match to the vanB targeting primers. We compensated for the loss of signal due to imperfect primer binding sites by increasing the *vanB* strain concentration in the contrived samples five-fold to achieve a robust dilution series. The high LoD for *bla*_CTX_ was a result of elevated background signal for this gene in the lot of reagents used in this study. All the AMR marker assays had within-and between-run precision of 100%.

The inclusivity of the assays was 100% for all alleles except *bla*_CTX_. Inclusivity for *bla*_CTX_ was limited to 54% because the assay has primers with perfect homology only to a single clade of *bla*_CTX-M_ variants (*bla*_CTX-M-3_ clade), but not the other highly divergent clades. However, based on the prevalence of *bla*_CTX_ variants in North America we estimated the capture of 67-75% of these variants in our assay (24). The exclusivity was estimated to be 100% for all markers.

Aside from being good laboratory practice, it is now a CAP requirement (CAP checklist item MIC.21855) to link AMR determinants to a specific organism in the final patient report (22). Linkage of the AMR marker with a specific bacterium has been challenging for culture-independent mNGS assays. For each AMR marker gene, our assays identify all the bacteria detected by the Karius Test known to carry that gene, referred to here as “carriers,” and their respective MPM values. A statistical model evaluates the likelihood of all the possible combinations of carriers and AMR gene copy number given an established prior to output a probability of linkage of AMR to each carrier. If the linkage cannot be established the result is reported as indeterminate.

Unlike many molecular and phenotypic assays, our assays not only detect *vanA*, *vanB, mecA*, and *mecC* but also differentiate between them. While *vanA* encodes resistance to vancomycin and other glycopeptides, *vanB* encodes resistance to vancomycin only (35). Although both *mecA* and *mecC* encode resistance to methicillin, *mecC* is much less common and more difficult to detect phenotypically, and as a result may be missed and reported as MSSA (36). In addition, this capability may increase the understanding of the prevalence of *mecC*. However, neither *vanB* nor *mecC* genes were detected in the clinical validation samples.

The Karius Test AMR marker detection covers 8 of 10 bacterial species driving global AMR-attributable deaths (1) and all of the ESKAPE pathogens which are important drivers of multidrug resistance (37), and the panel of 7 markers detect resistance to 5 different classes of antimicrobials, including anti-staphylococcal semisynthetic penicillins, glycopeptides, oxyimino-cephalosporins, aztreonam and carbapenems. Resistance to these drugs is among CDC’s top urgent and serious AMR threats in the US. In addition, we estimate from our commercial sample database that the target bacteria comprise approximately 18% of all reported detections and thus could be eligible for AMR marker detection.

Clinical accuracy was assessed with 115 plasma samples obtained from patients at 7 study sites with concordant culture results for any of the 18 target bacteria isolated from a variety of specimen types and correlated with available orthogonal phenotypic antimicrobial susceptibility test results in all cases and genotypic results when available. The performance characteristics of the SCC*mec* and *mecA* assays were assessed individually with the same 58 *S. aureus* detections and were found to be virtually identical except for DY which increased from 53.4% to 60.3% with the addition of the *mecA* assay. The performance for the combination of SCC*mec* and *mecA* for all staphylococci as they were intended to be used together in the AMR workflow (Fig. 1) showed excellent PPA, NPA, OPA, all >95%, and a DY of 70.0%. Although the number of samples that contained vancomycin-resistant enterococci was small, the PPA, NPA, and OPAwere 100%, and the DY was 83.3%.

The estimate of the PPA for the *bla*_CTX-M_ was only 55.5% based on the initial criteria of resistance to any oxyimino-beta-lactams, or aztreonam to label samples as putative ESBL producers. However, using susceptibility to cefepime as the sole criterion for labeling samples the PPA and DY improved to 83.3% and 97.1%, respectively.

In a recent comprehensive study of prevalence of carbapenem non-susceptible *Enterobacterales* (CNSE), 281 (62.4%) isolates carried *bla*_KPC_. OXA-48-like and metallo-β-lactamase (MBL) encoding genes were detected among seven (1.7%) and 12 (2.7%) isolates, respectively. One hundred and sixty-nine (37.6%) isolates did not produce carbapenemases (38). Given that only two of the samples included in clinical validation were phenotypically resistant to carbapenems, it is not surprising that we did not detect *bla*_KPC_.

Membrane permeability, together with derepression of the intrinsic beta-lactamase gene, is the global prevailing mechanism of carbapenem resistance in *P. aeruginosa*, but the acquired genes for carbapenemases are of increasing concern. Production of serine carbapenemases (e.g., KPC) by *Pseudomonas aeruginosa* are rare but MBLs may be present (39). The failure to detect a *bla*_KPC_ positive strain is likely due to the small sample size, rarity of the gene, and the diversity of mechanisms of carbapenem resistance in gram-negative bacilli.

A limitation of the study is the relatively small number of samples available for clinical validation. This was due to the stringent criteria used to qualify the samples: residual patient plasma samples which had one of target pathogens detected in the Karius Test, concordant culture results, applicable orthogonal phenotypic AST results, and adequate residual plasma volume. However, the supporting data and samples were collected from 7 study sites across the US and included a wide variety of sample types. Taken together, the results with this sample set suggest that they are generalizable and demonstrate that Karius Test can detect AMR markers in bacteria causing infections throughout the body. Although *mecC*, *vanB* and *bla*_KPC_ were not detected in our clinical validation samples, the excellent inclusivity, exclusivity, LoD, and precision data for these genes give us confidence that clinical performance for these makers should be similar to the more common markers detected in the clinical samples.

Another limitation is that the orthogonal phenotypic AST was performed with the Vitek2 and MicroScan Walkaway systems which are not standard reference methods. However, these systems are among the most widely used systems by clinical microbiology laboratories and despite their limitations may provide a better estimate of the performance of these assays in clinical practice.

We defined DY for each AMR marker as the percentage of tests that yielded a clinically actionable result (detected/not detected) and ranged from 56.8% for *bla*_KPC_ to 83.3% for *vanA*. While some might consider the DY low, the assays were optimized more accuracy than DY. This was best demonstrated by our results for methicillin resistance in staphylococci and cefepime resistance in gram-negative bacilli in which the OPA were estimated to be 95.2% and 97.1%, respectively.

Based on earlier identification of the cause of infection and increased diagnostic yield, mcfDNA sequencing results led to a change in antimicrobial management in 32%-72% of the reported cases (16, 17, 18, 19, 20). The addition of specific AMR markers to the Karius Test will provide clinicians more opportunities for optimization of therapy and anticipate the greatest clinical impact in those patients with infections in which the microbe is detected only by mcfDNA sequencing and in immunocompromised hosts who are particularly vulnerable to infections with multidrug-resistant bacteria. Faster identification of drug-resistant organisms has the potential to prevent the spread of multidrug resistance, to improve patient outcomes by enabling appropriate therapy to get patients on targeted therapy faster, and potentially to reduce mortality and hospitalization costs. In the AMR workflow we described, the results can be reported within a clinically relevant time frame with results being available 1 to 3 days after receipt of the sample in the Karius laboratory.

In summary, we report the validation of a novel application of mcfDNA sequencing for detection of AMR markers that has been incorporated into the Karius Test workflow without the need for a positive culture or intact microbial genomes. The key to success was enrichment for the ultrashort and rare mcfDNA fragments prior to sequencing for identification of AMR markers. The workflow could readily be adapted to expand the number of target bacteria and AMR genes as needed. Future studies will be focused on real-world evidence assessing the clinical impact of the Karius Test with expanded AMR marker detection since its launch.

### Data availability

The Karius Test is a proprietary laboratory-developed test, and thus there are several limitations on data availability that we declare here. Karius is unable to share the raw sequencing data underlying the test results evaluated here due to issues relating to patient privacy and consent. Additionally, the methodological details that can be provided to others to replicate the findings are limited due to intellectual property concerns. Taking these two limitations into account, summarized data used to reach the conclusions in this study are included in the manuscript and supplemental material.

## SUPPLEMENTAL MATERIAL

Supplemental Methods, Tables 1 to 6, and Figures 1 to 3.

Supplemental Table 7. All data for clinical validation samples (Excel workbook).

## ACKNOWLEDGMENTS

We acknowledge the support of the Karius cross-functional AMR core team in defining the scope, development, validation, and commercial launch of this new capability of the Karius Test. We thank the talented, experienced, and dedicated clinical laboratory operations and analytics teams at Karius for generating the test results and sequencing analyses, respectively, that were foundational to this study. We are also grateful for the thorough review by Karius’s Scientific Review Committee of this manuscript.

The following authors declare a conflict of interest: All employees of Karius, which markets the commercial test evaluated in this study, shared responsibilities for study design, analysis of the data, and writing and editing of the manuscript. John Joseph Farrell (OSF Healthcare), Sanjeet Dadwal, (City of Hope), Amy L. Carr, (AdventHealth Orlando), Arryn Craney (Orlando Health), Matthew Pike (Carle Foundation Hospital), James B. Wood (Indiana University School of Medicine) received grant support from Karius to support the collection and curation of orthogonal data that were fundamental to the clinical validation and reviewed the manuscript. This work was supported solely by funding from Karius.

