## Supplemental Methods, Tables 1 to 6, and Figures 1 to 3 for "Analytical and Clinical Validation of Direct Detection of Antimicrobial Resistance Markers by Plasma Microbial Cell-free DNA Sequencing"

### SUPPLEMENTAL MATERIAL

#### Methods

**Robustness of limit of detection to AMR gene copy number.** To examine the robustness of the limit of detection (LoD) to the copy number of the AMR gene, the copy number of the AMR genes was estimated using the Karius Test sequencing data (without targeting) for 100,000 MPM of contrived sheared pathogen genome carrying the AMR gene. The copy number was estimated using known ratios between the AMR gene and the pathogen genome length, the Karius Test estimated pathogen read count, and the AMR read counts from a custom alignment to the AMR genes (Supplemental Table 2). The observed AMR gene EDT was scaled by the copy number, e.g., with five gene copies, 20 EDT becomes 4 EDT, and recalculated the LoD. Each gene copy was assumed to contribute the same EDT signal as all the others, which is a conservative assumption, given that it does not allow for attribution of **most of** the EDT signal to a single gene copy.

**Precision. SCC*mec*.** To assess the precision of the SCC*mec* detection capabilities, four commercial (2 MRSA and 2 MSSA) samples were chosen for which there was an *S. aureus* call with a concentration that exceeded the minimum LoD at the QC minimum for unique WINC molecules (25,000). The expectation was that the call in all reruns of these samples would be identical to that of the original result. Each sample was requeued 3 times, either within the same sequencing batch (repeatability) or on different batches (reproducibility). **AMR gene panel.** To assess repeatability and within-lab reproducibility, samples from the LoD experiment were used. To assess precision of presence calls per gene, replicates containing the gene at an empirically tested concentration closest to, but not below, twice the presence LoD were chosen for a maximum total of 20 replicates examined per gene. The same procedure was repeated to assess precision of absence calls using empirically tested concentration closest to, but not below, twice the absence LoD. To evaluate repeatability and within-lab reproducibility in clinical samples, four samples were identified as MRSA by the Karius Test SCC*mec* caller where the *S. aureus* call is above twice the LoD for *mecA* and *mecC* detection. Six samples were identified following the same protocol for MSSA, with the *S. aureus* call above twice the LoD for determining *mecA* and *mecC* absence. The MRSA and MSSA plasma samples were made into two standardized clinical plasma pools. Three aliquots each from the MRSA and MSSA pools were tested within a sequencing run to determine repeatability. Three aliquots each from the MRSA and MSSA pools were tested across three sequencing runs to determine within-lab reproducibility.

**Housekeeping gene (HK) inclusivity/exclusivity.** Housekeeping gene targets for *S. aureus*, *S. epidermidis*, and *E. faecium* are used in the targeted AMR gene assay as a secondary check for species presence and judgment of AMR presence/absence. If the ratio of AMR EDT to housekeeping gene EDT for one of the three species departs from the expected relationship, the AMR call is "indeterminate." For example, lack of adequate housekeeping gene EDT would lower confidence in an AMR absence call, making it indeterminate. Housekeeping genes were chosen from species-specific core protein coding genes ([MetaPhlAn4 – the huttenhower lab. https://huttenhower.sph.harvard.edu/metaphlan/](https://huttenhower.sph.harvard.edu/metaphlan/) ) and filtered to ones where the nucleotide sequence would not cross-react with genomes of other pathogens. Only housekeeping genes for *S. aureus*, *S. epidermidis*, and *E. faecium* showed high species specificity and were then incorporated into the targeted AMR gene assay.

Housekeeping genes were checked against the Karius Test pathogen reference genome database and evaluated for the percent identity, alignment length, and number of species reference genome assemblies they aligned to. An ideal species-specific housekeeping gene would align fully at 100% identity to all of the species reference genomes in the database, but no other genomes.

Except for C-SA, all the chosen housekeeping genes align fully with almost 100% identity to all the available target species assemblies in the database (Supplemental Table 5). C-SA is one of 9 housekeeping genes for *S. aureus* and its presence or absence does not strongly bias the total housekeeping EDT recovery.

The cross-reactivity of the housekeeping genes with other species is very limited (Supplemental Table 6), the matches are either low percent identity (<80%), which would lead to a poor match of primers to the cross-reactive sequence or filtering via the analytical pipeline or the matches are very short DNA sequences (<10% of housekeeping gene length), which are unlikely to be bound by primers or contribute enough EDT signal to systematically bias housekeeping gene recovery.

### Tables

| **Supplemental Table 1.** *Staphylococcus aureus* genomes used for SCC*mec* LoD assessment. | | | | | | |
|---|---|---|---|---|---|---|
| **Assembly accession** | **MRSA/MSSA** | **Source** | **Geolocation** | **Collection Year** | **Strain** | **SCC*mec* subtype**[a] |
| GCF_001018905.2 | MRSA | Wound | USA:Washington | - | NRS387; USA800; 1045 (FDAARGOS_26) | IVc(2B) / IV |
| GCF_001018875.2 | MRSA | Sputum | USA:Tennessee | - | NRS127 (FDAARGOS_22) | II(2A) / II |
| GCF_001046135.1 | MRSA | Blood | USA: New York | 2009 | C1440 t008 | IVa(2B) / - |

| | | | | | | |
|---|---|---|---|---|---|---|
| GCF_001046615.1 | MRSA | Nares | USA: Florida | 2009 | C1555, t008 | IVg(2B) / - |
| GCF_001019565.2 | MRSA | Blood | Denmark | 1964 | E2125 (FDAARGOS_46) | I(1B) / I |
| GCF_001018685.2 | MSSA | Buttock abscess | USA:Texas | 2002 | TCH 959 (USA300-HOU-MS); ATCC BAA-1718 (FDAARGOS_4) | None |
| GCF_001018975.2 | MSSA | - | - | - | NRS143 (FDAARGOS_30) | None |

| | | | | | | |
|---|---|---|---|---|---|---|
| GCF_001018715.2 | MSSA | - | - | - | NRS128 (FDAARGOS_11) | None |
| GCF_001019375.2 | MSSA | Vaginal tampon | - | - | NRS156 (FDAARGOS_17) | None |
| GCF_001018725.2 | MSSA | - | French Guiana | 1996 | NRS129 (FDAARGOS_10) | None |

[a]SCC*mec*Finder inferred / BioSample metadata

https://github.com/ajaybabu27/sccmecfinder.

| **Supplemental Table 2.** Organisms and genomes used to establish LoD. | | | | |
|---|---|---|---|---|
| **Organism** | **Resistance Genotype** | **Estimated gene copy number** | **ATCC Strain ID (Strain Name)** | **Mix** |
| *Staphylococcus aureus* | *mecA*(+) | 1 | 700699 (*S. aureus subsp. aureus* Mu50) | P3.2 |
| *S. aureus* | *mecC*(+) | 1 | BAA-2313 (*S. aureus* M10/0148) | P3.3 |
| *Enterococcus faecium* | *vanA*(+) | 6 | 700221 (*E. faecium* VRE) | P3.2 |
| *E. faecalis* | *vanB*(+) | 2 | 700802 (*E. faecalis* V583) | P3.3 |
| *Klebsiella pneumoniae* | $bla_{KPC}$(+) | 3 | BAA-1705 (*K. pneumoniae* ART 2008133) | P3.3 |
| *Escherichia coli* | $bla_{CTX-M}$(+) | 2 | BAA-2326 (*E. coli* TY-2482) | P3.2 |

| **Supplemental Table 3.** Limit of Detection experimental design (N = 260). | |
|---|---|
| **Dilution** | **Pathogen Input Concentration in MPM* (No. replicates)** |
| 1 (P3.2, P3.3) N=20 | 100,000 (x10) **vanB*: 500,000 |
| 2 (P3.2, P3.3) N=20 | 31,600 (x10) **vanB*: 158,000 |
| 3 (P3.2, P3.3) N=40 | 10,000 (x20) **vanB*: 50,000 |
| 4 (P3.2, P3.3) N=40 | 3,160 (x20) **vanB*: 15,800 |
| 5 (P3.2, P3.3) N=40 | 1,000 (x20) **vanB*: 5,000 |
| 6 (P3.2, P3.3) N=40 | 316 (x20) **vanB*: 1,580 |
| 7 (P3.2, P3.3) N=40 | 100 (x20) **vanB*: 500 |

| | |
|---|---|
| 8 (P3.2, P3.3) N=20 | 32 (x10) *vanB*: 160 |
| 9 (plasma background) | 0 (x40 or 4 per sequencing run) |

| **Supplemental Table 4.** LoD per target and copy number for the AMR gene panel. | | |
|---|---|---|
| **Target** | **Original LoD** | **LoD at 1 gene copy** |
| *mecA* presence | 662 MPM | 662 MPM |
| *mecC* presence | 808 MPM | 808 MPM |
| *vanA* presence | 425 MPM | 3,296 MPM |
| *vanB presence* | *4,985 MPM* | *6,012 MPM* |
| *bla*$_{CTX-M}$ presence | 6,107 MPM | 1,458 MPM |
| *bla*$_{KPC}$ presence | 1,346 MPM | 3,169 MPM |

| **Supplemental Table 5.** HK gene inclusivity | | | | |
|---|---|---|---|---|
| **Species** | **HK gene name** | **% matching assemblies** | **Median alignment coverage** | **Median percent identity** |
| *S. aureus* | *A-SA* | 22/22 (100%) | 1.000000 | 99.4445 |
| | *B-SA* | 22/22 (100%) | 1.000000 | 97.8210 |
| | *C-SA* | 13/22 (59%) | 1.000000 | 99.4410 |
| | *D-SA* | 22/22 (100%) | 1.000000 | 98.6670 |
| | *E-SA* | 22/22 (100%) | 1.000000 | 99.0820 |
| | *glpF* | 22/22 (100%) | 1.000000 | 99.6775 |
| | *orfX* | 22/22 (100%) | 1.000000 | 98.5420 |
| | *arcC* | 22/22 (100%) | 1.000000 | 99.3420 |
| | *aroE* | 22/22 (100%) | 1.000000 | 98.9040 |
| *S. epidermidis* | *A-SE* | 8/8 (100%) | 1.000000 | 98.9820 |

| | | | | |
|---|---|---|---|---|
| | *B-SE* | 8/8 (100%) | 1.000000 | 98.2330 |
| | *C-SE* | 8/8 (100%) | 1.000000 | 97.8850 |
| | *D-SE* | 8/8 (100%) | 1.000000 | 96.9095 |
| | *yqiL* | 8/8 (100%) | 1.000000 | 99.1545 |
| | *pyrR* | 8/8 (100%) | 1.000000 | 97.1960 |
| | *aroE* | 8/8 (100%) | 0.998960 | 97.8145 |
| | *tpiA* | 8/8 (100%) | 1.000000 | 99.4090 |
| *E. faecium* | *atpA* | 20/20 (100%) | 1.000000 | 97.8420 |
| | *purK* | 20/20 (100%) | 0.998984 | 99.7965 |
| | *ddl* | 20/20 (100%) | 1.000000 | 98.7100 |

| **Supplemental Table 6.** HK gene exclusivity | | | | |
|---|---|---|---|---|
| **Species** | **HK gene name** | **no. matching assemblies of non-target species** | **Median alignment coverage** | **Median percent identity** |
| *S. aureus* | *A-SA* | – | – | – |
| | *B-SA* | – | – | – |
| | *C-SA* | 1 | 0.061453 | 93.9390 |
| | *D-SA* | 4 | 0.104000 | 89.7440 |
| | *E-SA* | – | – | – |
| | *glpF* | 52 | 1.000000 | 78.3550 |
| | *orfX* | 110 | 0.995833 | 79.7920 |
| | *arcC* | 45 | 0.574561 | 72.5000 |
| | *aroE* | 36 | 0.995614 | 74.8565 |

| | | | | |
|---|---|---|---|---|
| *S. epidermidis* | *A-SE* | – | – | – |
| | *B-SE* | 1 | 0.037691 | 93.7500 |
| | *C-SE* | 1 | 0.069374 | 87.8050 |
| | *D-SE* | 1 | 0.033435 | 93.9390 |
| | *ymaiL* | 8 | 0.350242 | 72.2390 |
| | *pyrR* | 57 | 0.271028 | 75.7510 |
| | *aroE* | 42 | 0.979210 | 72.4470 |
| | *tpiA* | 887 | 0.891253 | 72.4600 |
| *E. faecium* | *atpA* | 1185 | 0.953237 | 75.7125 |
| | *purK* | 20 | 0.887195 | 75.9730 |
| | *ddl* | 74 | 0.969892 | 79.3790 |

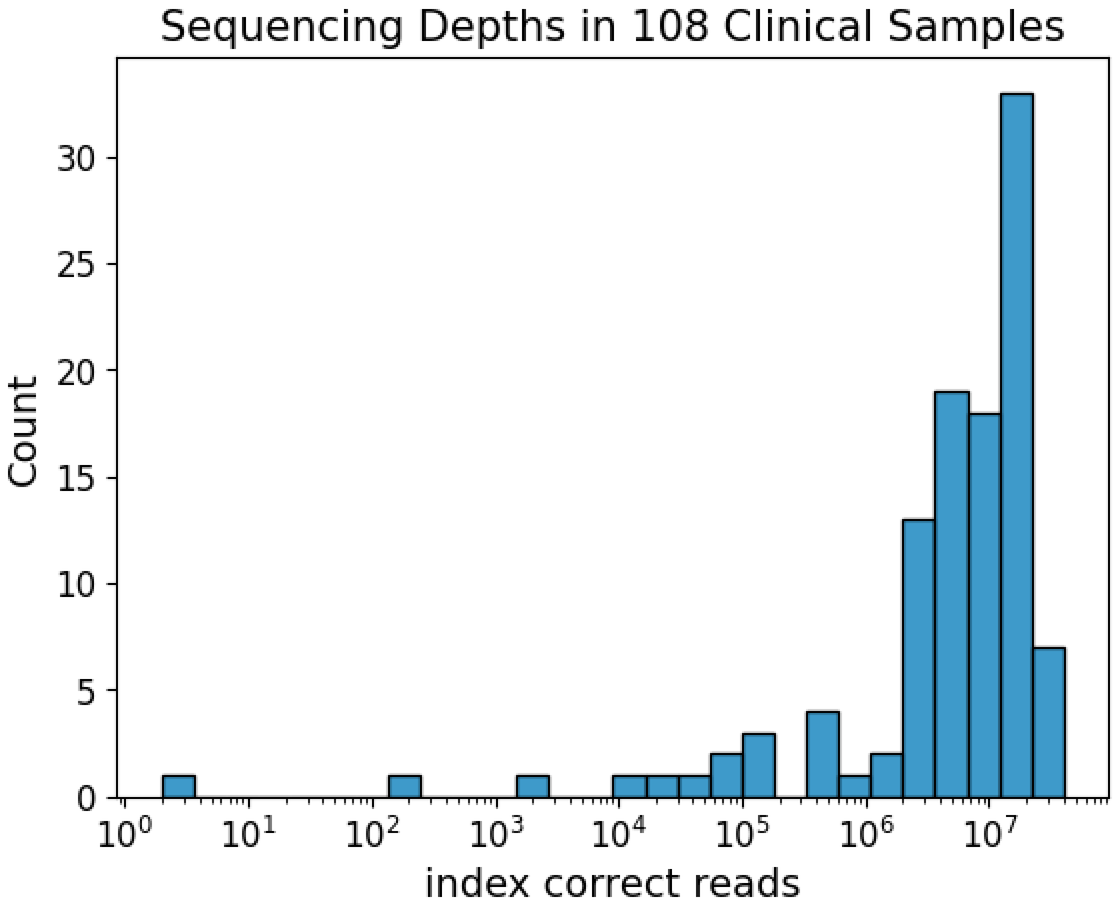


### Figures

**Supplemental Fig. 1.** Distribution of index correct read counts across 108 clinical samples.

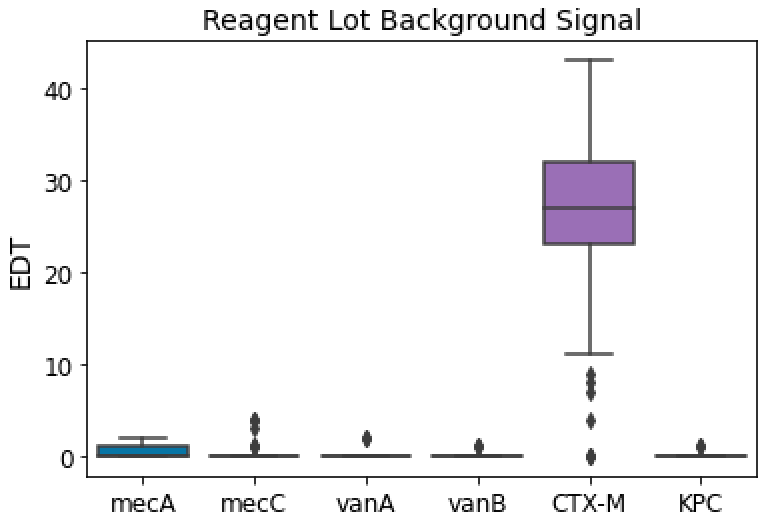


**Supplemental Fig. 2.** Distribution of EDTs in negative control samples for each AMR target gene.

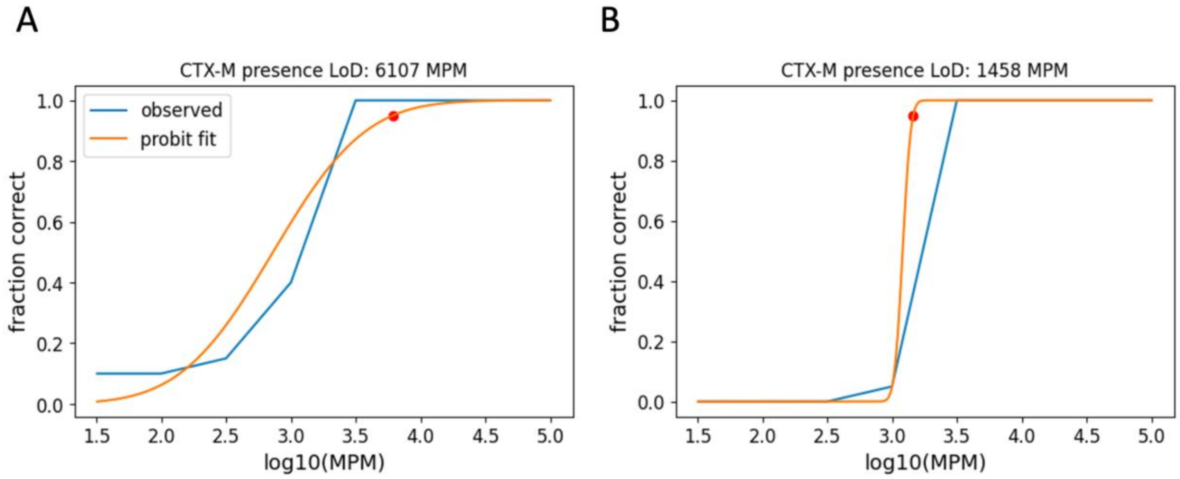


**Supplemental Fig. 3.** Probit fits for estimating CTX-M presence LoD using the (A) original dilution series and (B) simulated dilution series assuming one CTX-M gene copy. Orange lines reflect the probit model fit and blue lines reflect the observed data of fraction positive calls per concentration.
